# High resolution imaging and five-year tuberculosis contact outcomes

**DOI:** 10.1101/2023.07.03.23292111

**Authors:** Hanif Esmail, Anna K Coussens, Friedrich Thienemann, Bianca Sossen, Sandra L Mukasa, James Warwick, Rene T. Goliath, Nashreen Omar Davies, Emily Douglass, Amanda Jackson, Francisco Lakay, Elizabeth Streicher, Jacob E. Munro, Marilou H Barrios, Torben Heinsohn, Liana Macpherson, Dylan Sheerin, Saalikha Aziz, Keboile Serole, Remy Daroowala, Arshad Taliep, Petri Ahlers, Stephanus T. Malherbe, Rory Bowden, Robin Warren, Gerhard Walzl, Laura E. Via, Melanie Bahlo, Karen R. Jacobson, C. Robert Horsburgh, Padmini Salgame, David Alland, Clifton Earl Barry, JoAnne L. Flynn, Jerrold J Ellner, Robert J Wilkinson

**Author notes:** equal contribution (Hanif Esmail, Anna K Coussens). Corresponding author: Anna Coussens.

## Abstract

**Background:** The evolution of tuberculosis (TB) disease during the clinical latency period remains incompletely understood.

**Methods:** 250 HIV-uninfected, adult household contacts of rifampicin-resistant TB with a negative symptom screen underwent baseline ^18^F-Fluorodeoxyglucose positron emission and computed tomography (PET/CT), repeated in 112 after 5-15 months. Following South African and WHO guidelines, participants did not receive preventive therapy. All participants had intensive baseline screening with spontaneous, followed by induced, sputum sampling and were then observed for an average of 4.7 years for culture-positive disease. Baseline PET/CT abnormalities were evaluated in relation to culture-positive disease.

**Results:** At baseline, 59 (23.6%) participants had lung PET/CT findings consistent with TB of which 29 (11.6%) were defined as Subclinical TB, and 30 (12%) Subclinical TB-inactive. A further 83 (33.2%) had other lung parenchymal abnormalities and 108 (43.2%) had normal lungs. Over 1107-person years of follow-up 14 cases of culture-positive TB were diagnosed. Six cases were detected by intensive baseline screening, all would have been missed by the South African symptom-based screening strategy and only one detected by a WHO-recommended chest X-Ray screening strategy. Those with baseline Subclinical TB lesions on PET/CT were significantly more likely to be diagnosed with culture-positive TB over the study period, compared to those with normal lung parenchyma (10/29 [34.5%] vs 2/108 [1.9%], Hazard Ratio 22.37 [4.89-102.47, p<0.001]).

**Conclusions:** These findings challenge the latent/active TB paradigm demonstrating that subclinical disease exists up to 4 years prior to microbiological detection and/or symptom onset. There are important implications for screening and management of TB.

## Introduction

One quarter of the world’s population is considered to have latent tuberculosis (TB) infection (i.e., evidence of immune sensitization in the absence of clinical signs or symptoms of disease).^1^ The risk of developing disease is elevated for 5-10 years following exposure.^2–4^ Conventionally, latent TB is conceived as a state in which *Mycobacterium tuberculosis (Mtb)* is successfully contained within microscopic granulomas which limit replication, preventing invasive pathology, symptoms and infectiousness.^5, 6^ Active TB disease is considered to arise when granulomatous control fails leading to evident, invasive pathology associated with symptoms, detectable bacilli and infect**i**ousness.^7^ This paradigm strongly influences programmatic management.^8, 9^

Insight into pathological events during the period of clinical latency is limited. A more nuanced concept of disease evolution that encompasses heterogeneity amongst persons with immune sensitisation with some having evidence of disease in the absence of symptoms has been proposed.^10, 11^ Early lung disease is well-described in autopsy studies and animal models and is characterized by a cellular infiltrate spreading bronchogenically.^12–14^ In a previous study we showed that high-resolution anatomical and functional imaging by ^18^F-Fluorodeoxyglucose combined positron emission and computed tomography (PET/CT) was highly sensitive to identify pathology consistent with TB disease in asymptomatic HIV-infected, individuals considered to have latent TB, defining an imaging phenotype for subclinical disease.^15^

To further understand TB progression, the aims of this study were to (1) determine the proportion of HIV-uninfected, symptom-free, TB household contacts (HHC) with baseline PET/CT abnormalities suggestive of subclinical TB disease and (2) compare the proportion with and without these abnormalities diagnosed with culture positive TB following intensive respiratory sampling and close follow-up over 5 years without preventive treatment.

## Methods

### Study design and oversight

A prospective, observational, cohort study was conducted in HIV-uninfected, adult HHC of rifampicin-resistant (RR) TB cases who, consistent with national and international guidelines, were not provided preventive therapy.^16, 17^ The study was approved by the human research ethics committees of the University of Cape Town (HREC 449/2014), Boston University (H-35831), Rutgers University (Pro2018001966), and the Division of Microbiology and Infectious Diseases of NIH (DMID 16-0112). This report follows STROBE guidelines for cohort studies.

### Recruitment of participants

Recruitment was between November 2014 and September 2017 with follow-up until May 2021. All participants were residents of Khayelitsha, South Africa. Index cases (>15 years old) with RR pulmonary TB consented to a household visit. Adult HHC not on TB treatment were baseline screened following initial consent. Participants further consented to PET/CT imaging and blood sampling (**Supplementary Methods**) if not meeting the following exclusion criteria; HIV infection, symptoms of TB by South African national guidelines (2 weeks cough, fever, weight loss, night sweats^17^), clinical signs of TB, acute illness, age >65 years, smoker >30 pack-years, malignancy, chronic lung infection or inflammation, inhaled or systemic steroid use within previous 2 weeks, breast-feeding, pregnant, planning pregnancy, unable to be followed up, uncontrolled diabetes mellitus, unwilling, or at investigator discretion (**Figure 1**).

**Figure 1.**
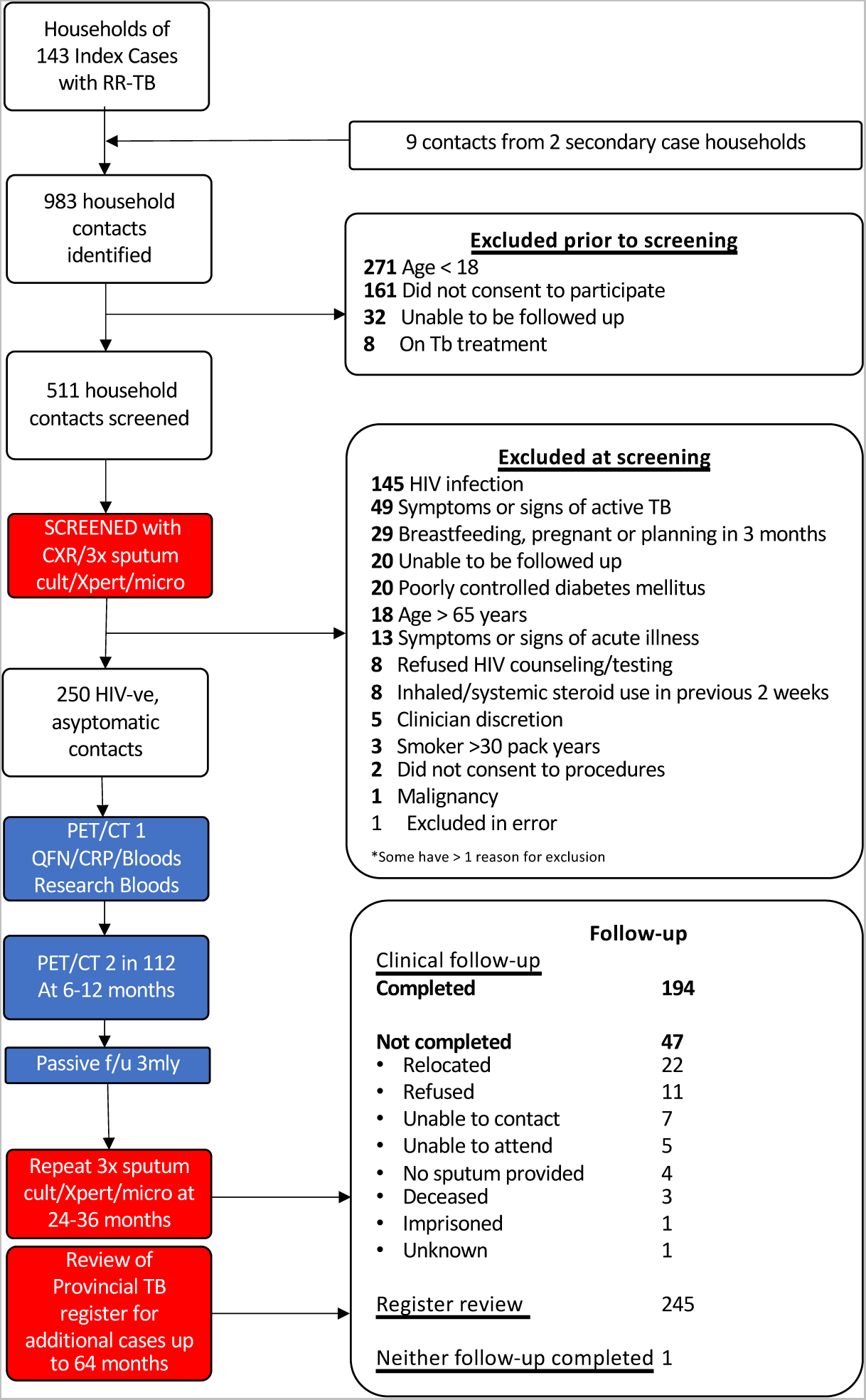
Study flow diagram. Shows number of individuals screened, excluded, consented and followed up. Cult= culture, micro=microscopy

### Investigations to diagnose TB

At baseline all participants underwent TB symptom screening, digital chest radiography (CXR). All were invited to produce a sputum sample spontaneously if able (irrespective of symptom screen and CXR status) and then underwent further sputum induction using hypertonic saline until three sputum samples were obtained. All sputum samples were sent for smear, Xpert MTB/RIF and bacterial (MGIT) culture (**Supplementary Methods**). Follow-up was both active and passive. Repeat PET/CT was performed at 5-15 months in 112/250 participants and bronchoscopy and lavage culture performed on 28/112 (**Supplementary Methods**). All participants were actively screened for TB between 23 to 38 months with three sputum samples (induced if needed) and any time before this visit, if they reported TB symptoms during 3-montly phone calls. The Western Cape Provincial Health Data Centre (PHDC) database was cross-checked for additional cases in May 2021.^18^

Participants were considered to have culture-confirmed TB if any specimen was *Mtb* positive by culture. Participants were considered to have clinically confirmed TB if there was a decision to start TB treatment made by statutory services without a positive culture.

### PET/CT classification

Two readers provided independent structured reports, differences were resolved by a third reader. All were blinded to clinical history and microbiological results (**Supplementary Methods**). Baseline scans were classified into four mutually exclusive categories based on previous work^15^:

● <u>Radiographically consistent with TB disease (Subclinical TB):</u> Presence of infiltrate and/or FDG-avid nodule(s) within the upper lobes or apical segment of lower lobes.
● <u>Radiographically consistent with inactive TB (Subclinical TB-inactive):</u> Presence of fibrotic scar(s) within the upper lobes or apical segment of lower lobes.
● Other parenchymal abnormalities not meeting definitions above.
● Normal lung parenchyma.

### Sample size

Sample size was governed by the expected proportion with PET/CT parenchymal abnormalities consistent with Subclinical TB or Subclinical TB-inactive. We predicted 50% would become infected following exposure, of whom 30-40% would have abnormalities based on prior experience.^15^ We therefore expected 15-20% to have PET/CT abnormalities with a sample of 250 providing precision of approximately ±5% with 95% confidence.

### Consideration for different screening strategies

As the intensity of baseline screening could affect the likelihood of culture positive cases being identified during follow-up, to help interpret the findings within a real-world context, we considered the culture positive events in relation to 3 baseline screening approaches of varying stringency.

- <u>Study screening approach:</u> 3 x sputum culture, induced if needed, irrespective of symptom screen or CXR findings.
- <u>South African national guidelines:</u> Spontaneous sputum Xpert investigation at baseline only if symptom screen positive.
- <u>WHO systematic screening guidelines (2021):</u> Spontaneous sputum Xpert investigation at baseline only if symptom screen positive or CXR suggestive of TB.

### Statistical analysis

Statistical analysis was conducted in Stata ver. 17.0 (StataCorp). Non-parametric data was compared using Mann-Whitney *U* test or Kruskal Wallis with Dunn’s *post hoc* test and parametric data compared using t-test or ANOVA. Proportions were compared by χ^2^ or Fisher’s exact test. We used Cox proportional-hazards regression to assess the impact of PET/CT classification on time to culture positivity, the proportional hazards assumption was assessed by Schoenfeld residuals. Multivariate modelling was undertaken to assess the impact of covariates (previous TB, age, sex, QuantiFERON status and hours of contact/day with index) by forward selection and assessment by Likelihood Ratio test. Only participants with complete data for these variables were included (247 participants).

## Results

### Recruitment

983 household members of 145 index cases with RR-TB were identified, of whom 271 (28%) were <18 years old. 511 adult participants were screened for eligibility. 261 (51%) participants were excluded including 145 with HIV infection and 49 TB symptom positive (**Figure 1**).

### Clinical characteristics

250 HIV uninfected participants were enrolled: median age 30 years (IQR 23-43), 60% women. Thirty-four (13.6%) had a previous TB history (median 10 years prior (IQR 7-24). All participants had a negative symptom screen. 248 had a valid QuantiFERON-TB Gold (QFT-G) result and 241 a QuantiFERON-TB Plus (QFT-Plus) result. 205 (82%) were positive by at least one test, thus defined as having latent TB infection. 249 participants underwent CXR, 37 (14.9%) had changes suggestive of TB (**Table 1**). 160 (64%) produced a sputum sample spontaneously and 249 following induction (237 (94.8%) provided a total of three samples). The cohort was followed for 1107 person-years (median 4.7 years). 194 (77.6%) had repeat sputa between 23-38 months and 247 (98.8%) were tracked through the PHDC. One participant was not followed up in either way.

**Table 1.** Baseline characteristics, clinical and radiographic findings, and TB outcomes of HHC who underwent PET/CT. Values are n (%) or median (IQR). Denominator values are indicated in the column descriptor, or indicated in a cell when values were missing. Relationship between PET/CT categories and baseline characteristics analysed for categorical variables analysed by χ^2^ or Fisher’s exact test, for numerical variables by Kruskal Wallis with *post hoc* analysis using Dunn’s multiple comparison testing. Bold and superscript number indicates which post-hoc numerical comparisons are significantly different (p<0.05): 1 = in comparison with no lung lesions, 2 = in comparison with Other lung lesions, 3 = in comparison with Subclinical-inactive TB. BMI, body mass index; Clinical TB, TB symptom positive *Mtb* culture and Xpert negative; CRP, C-reactive protein; CXR, chest X-ray; ESR, erythrocyte sedimentation rate; F, female; HU_max_, maximum Hounsfield units; IFNγ, interferon-gamma; LN, lymph node; Neut, neutrophil; N:L, neutrophil:lymphocyte ratio in blood; N:M, neutrophil:monocyte ratio in blood; QFN+, QuantiFERON positive; SUV_max_, maximum standardised uptake value; VS, visual score; WCC, whole cell count; Xpert, GeneXpert version 3.0: superscript 4 = detected at baseline, 5 = detected at follow-up.

| Variable | All HHC<br>n=250 | Subclinical TB<br>n=29 | Subclinical TB-Inactive<br>n=30 | Other lung lesions<br>n=83 | Normal lung<br>n=108 | p-value |
| --- | --- | --- | --- | --- | --- | --- |
| **Demographics** |  |  |  |  |  |  |
| Age (years) | 30 (23-43) | **43 (30-50)<sup>1,2</sup>** | **41 (28-54)<sup>1,2</sup>** | 28 (23-39) | 27 (22-40) | **<0.001** |
| Sex (Female) | 150 (60%) | 14 (48.3%) | 17 (56.7%) | 53 (63.9%) | 66 (61.1%) | 0.5 |
| Previous TB | 34 (13.6%) | 14 (48.3%) | 15 (50%) | 2 (2.4%) | 3 (2.8%) | **<0.001** |
| Daily index contact | 208 (83.2%) | 24 (82.8%) | 26 (86.7%) | 72 (86.7%) | 86 (79.6%) | 0.57 |
| >6hrs/day with index | 96/249 (38.5%) | 14 (48.3%) | 10 (33.3%) | 37 (44.6%) | 35/107 (32.7%) | 0.23 |
| 1st degree relative | 106 (42.4%) | 9 (31.0%) | 19 (63.3%) | 39 (47.0%) | 39 (36.1%) | 0.11 |
| Ever smoked | 85 (34%) | 12 (41.3%) | 15 (50%) | 22 (26.5%) | 36 (33.3%) | 0.10 |
| BMI | 28.2 (22.2-34) | **24.0 (19.8-29.3)<sup>2,3</sup>** | **25.5 (21.3-34.6)<sup>2,3</sup>** | 29.2 (22.37-33.49) | 28.5 (23.5-35.4) | **0.02** |
| **Clinical Investigations** |  |  |  |  |  |  |
| CXR TB Suggestive | 37/249 (14.9%) | 16 (55.2%) | 10/29 (34.5%) | 2 (2.4%) | 9 (8.3%) | **<0.001** |
| Proportion QFN+ Gold or Plus | 205/248 (82.7%) | 28 (96.6%) | 27 (90.0%) | 72 (86.7%) | 78/106 (73.6%) | **0.007** |
| QFN Nil IFNγ (IU/ml) | 0.03 (0.00-0.13) | 0.08 (0.01-0.14) | 0.01 (0.00-0.08) | 0.03 (0.00-0.18) | 0.03 (0.00-0.12) | 0.17 |
| QFN Gold Ag-Nil IFNγ (IU/ml) | 6.82 (0.66-41.83) | **14.62 (3.11-56.85)<sup>1</sup>** | **10.2 (1.4-57.0)<sup>1</sup>** | **7.6 (0.9-48.0)<sup>1</sup>** | 3.79 (0.1-19.1) | **0.01** |
| QFN Plus-1 Ag-Nil IFNγ (IU/ml) | 5.53 (0.45-29.57) | **12.90 (2.39-36.93)<sup>1</sup>** | **10.70 (1.95-34.69)<sup>1</sup>** | **5.53 (0.95-40.26)<sup>1</sup>** | 2.78 (0.15-16.57) | **0.02** |
| QFN Plus-2 Ag-Nil IFNγ (IU/ml) | 6.26 (0.59-33.37) | **18.01 (2.58-44.46)<sup>1</sup>** | **10.76 (1.80-33.72)<sup>1</sup>** | **6.49 (0.76-48.44)<sup>1</sup>** | 3.48 (0.24-20.50) | **0.02** |
| CRP (mg/L) | 3 (1-7) | 4 (1-11) | 3.5 (1-7) | 2.3 (1-5) | 2 (1-7) | 0.44 |
| CRP≥10 mg/L | 40/247 (16.2%) | 8 (27.6%) | 4 (13.3%) | 11/82 (13.4%) | 17/106 (16.0%) | 0.33 |
| ESR (mm/Hr) | 15 (5-30) | 20 (8-33) | 20.5 (9-31) | 11 (4-30) | 13.5 (3-26) | 0.10 |
| WCC (x10<sup>9</sup>/L) | 5.91 (4.88-7.54) | 7.04 (5.97-8.01) | 5.75 (5.06-7.18) | 5.78 (4.54-7.39) | 5.85 (4.74-7.62) | 0.06 |
| Neut (x10<sup>9</sup>/L) | 3.28 (2.27-4.54) | **4.00 (3.39-5.14)<sup>1,2,3</sup>** | 3.25 (2.33-4.46) | 2.94 (2.18-4.64) | 3.19 (2.25-4.42) | **0.04** |
| N:L | 1.59 (1.12-2.24) | **2.21 (1.60-2.67)<sup>1,2,3</sup>** | 1.56 (1.21-2.32) | 1.50 (1.10-2.22) | 1.60 (1.07-2.09) | **0.03** |
| N:M | 8.04 (6.21-10.53) | 9.17 (7.13-12.24) | 7.24 (5.81-10.47) | 8.15 (6.49-10.40) | 7.99 (6.11-10.16) | 0.24 |
| L:M | 5.09 (4.00-6.49) | 4.50 (3.47-5.85) | 5.31 (3.44-6.65) | 4.99 (4.31-6.77) | 5.15 (4.11-6.26) | 0.24 |
| **PET/CT Imaging Findings** |  |  |  |  |  |  |
| Lung total lesions | 1 (0-3) | **6 (4-10)<sup>1,2</sup>** | **5 (2-8)<sup>1,2</sup>** | 1 (1-3) | 0 (0-0) | **<0.001** |
| total infiltrates | 0 (0-0) | **1 (0-2)<sup>2,3</sup>** | 0 (0-0) | 0 (0-0) | NA | **<0.001** |
| total fibrotic scars | 0 (0-2) | **1 (0-2.5)<sup>2,3</sup>** | **2 (1-3)<sup>2</sup>** | 0 (0-0) | NA | **<0.001** |
| total nodules | 1 (0.75-3) | **3 (1-5)<sup>2</sup>** | 1 (0-4.25) | 1 (1-2) | NA | **0.028** |
| total cavities | 0 (0-0) | **0 (0-0.5)<sup>2,3</sup>** | 0 (0-0) | 0 (0-0) | NA | **<0.001** |
| Lung largest lesion size (mm) | 11.15 (4.82-32.55) | **44.19 (26.2-56.3)<sup>2</sup>** | **33.3 (26.5-45.7)<sup>2</sup>** | 5 (4-9.66) | NA | **<0.001** |
| Lung maximum VS | 0 (0-1) | **3 (2-3)<sup>2,3</sup>** | **0 (0-1)<sup>2</sup>** | 0 (0-0) | NA | **<0.001** |
| Lung SUV<sub>max</sub> | 1.29 (0.94-1.89) | **2.63 (1.77-5.77)<sup>2,3</sup>** | **1.45 (1.18-1.9)<sup>2</sup>** | 1.07 (0.81-1.35) | NA | **<0.001** |
| Lung HU<sub>max</sub> | 115 (-43-481) | **185 (88-648)<sup>2</sup>** | **670.5 (140-1123)<sup>2</sup>** | 7.5 (-129-179) | NA | **<0.001** |
| LN total lesions | 0 (0-2) | **2 (0-3)<sup>1,2</sup>** | **1.5 (0-3)<sup>1,2</sup>** | 0 (0-2) | 0 (0-1) | **<0.001** |
| FDG-avid LN present | 53 (21.2%) | 16 (55.2%) | 12 (40%) | 15 (18.07%) | 10 (9.3%) | **<0.001** |
| Largest abnormal LN (mm) | 8.5 (5.8-11.2) | 11.0 (8.35-13.4) | 8.0 (5.42-12.2) | 8.4 (5.0-10.3) | 8.0 (5.4-10.0) | 0.06 |
| LN maximum VS | 1.5 (1-3) | **3 (2-3)<sup>1,2,3</sup>** | **2 (1-3)<sup>1</sup>** | 1 (1-2) | 1 (1-2) | **0.005** |
| LN SUV<sub>max</sub> | 2.63 (1.90-4.10) | **3.51 (2.63-4.87)<sup>1,2</sup>** | 2.60 (2.19-3.53) | 2.25 (1.60-3.36) | 2.11 (1.74-3.46) | **0.02** |
| LN HU<sub>max</sub> | 549.5 (134.5-1041.5) | 274.5 (94.5-7685) | 786 (158-1092) | 488 (136-1013) | 658 (208-1115) | 0.25 |
| **TB Outcome** |  |  |  |  |  |  |
| Culture positive at baseline (treated) | 6 (2.4%) | 6 (20.7%) | 0 (0%) | 0 (0%) | 0 (0%) | **<0.001** |
| Culture positive at follow-up (treated) | 8/249 (3.2%) | 4 (18.8%) | 1 (3.3%) | 1 (1.2%) | 2/107 (1.9%) | **0.007** |
| Clinical TB at follow-up (treated) | 4/249 (1.6%) | 2 (6.9%) | 1 (3.3%) | 1 (1.2%) | 0/107 (0%) | **0.01** |
| Xpert positive culture negative (not treated, asymptomatic) | 4 (1.6%) | 1 (3.4%)<sup>5</sup> | 2 (6.7%)<sup>4,5</sup> | 0 (0%) | 1 (0.9%)<sup>4</sup> | **0.04** |
| Any TB or Mtb detected over study period | 22 (8.2%) | 13 (44.8%) | 4 (13.3%) | 2 (2.5%) | 3 (2.8%) | **<0.001** |
| Symptoms at Any treated TB diagnosis | 7/18 (38.9%) | 2/12 (16.7%) | 2/2 (100%) | 2/2 (100%) | 1/2 (50%) | **0.03** |
| Symptoms at culture positive TB diagnosis | 3/14 (21.4%) | 0/10 (0%) | 1/1 (100%) | 1/1 (100%) | 1/2 (50%) | **0.01** |

### Baseline PET/CT findings in asymptomatic household contacts

On baseline PET/CT imaging, 142 (56.8%) had abnormalities within the lung parenchyma: 29 (11.6%) Subclinical TB, 30 (12%) Subclinical TB-inactive, 83 (33.2%) other abnormalities. 108 (43.2%) had no lung abnormalities (**Table 1 and Supplementary Table S1**). 105 participants (42%) had mediastinal and hilar lymph node abnormalities, of whom 53 had FDG-avid lymph nodes (**Table 1 and detailed in Supplementary Appendix**). Those with Subclinical TB and Subclinical TB-inactive had greater total lymph node lesions and maximum visual score (≤0.005).

Approximately half (48.3-50%) of those categorised as Subclinical TB and Subclinical TB-inactive had a previous TB history, compared to 2.4-2.8% of those with normal or other lung lesions (p<0.001). Of the 216 with no history of TB, 15 (6.9%) had Subclinical TB and 15 (6.9%) Subclinical TB-inactive. Of these, 7/15 (46.7%) and 1/14 (7.1%; one no CXR), respectively, had a CXR reported as suggestive of TB (**Supplementary Table S2**).

### Risk of culture positivity in relation to baseline PET/CT findings

Over the study period 14/250 (5.6%) were diagnosed with culture positive TB (**Table 1**). Six were diagnosed following intensive baseline screening and eight during follow-up after a median of 32 months (IQR 9.5-37.5), of which only 3/8 (37.5%) were symptomatic at follow-up diagnosis (**Supplementary Table S3**). Of the six baseline culture positive participants, none would have been diagnosed if screened according to South African national guidelines and only 1 diagnosed if screened following WHO guidelines (**Figure 2**). Four additional participants were treated for TB during follow-up, two with positive Xpert alone, and two clinically diagnosed. A further four had a positive Xpert alone without symptoms and were not treated (with no progression observed during follow-up). These eight patients were not included in the primary analysis but in two sensitivity analyses (**Figure 3 and Supplementary Results-2.5**). Individual longitudinal imaging and clinical findings for all 22 participants are shown in **Supplementary Figure S1**.

**Figure 2.**
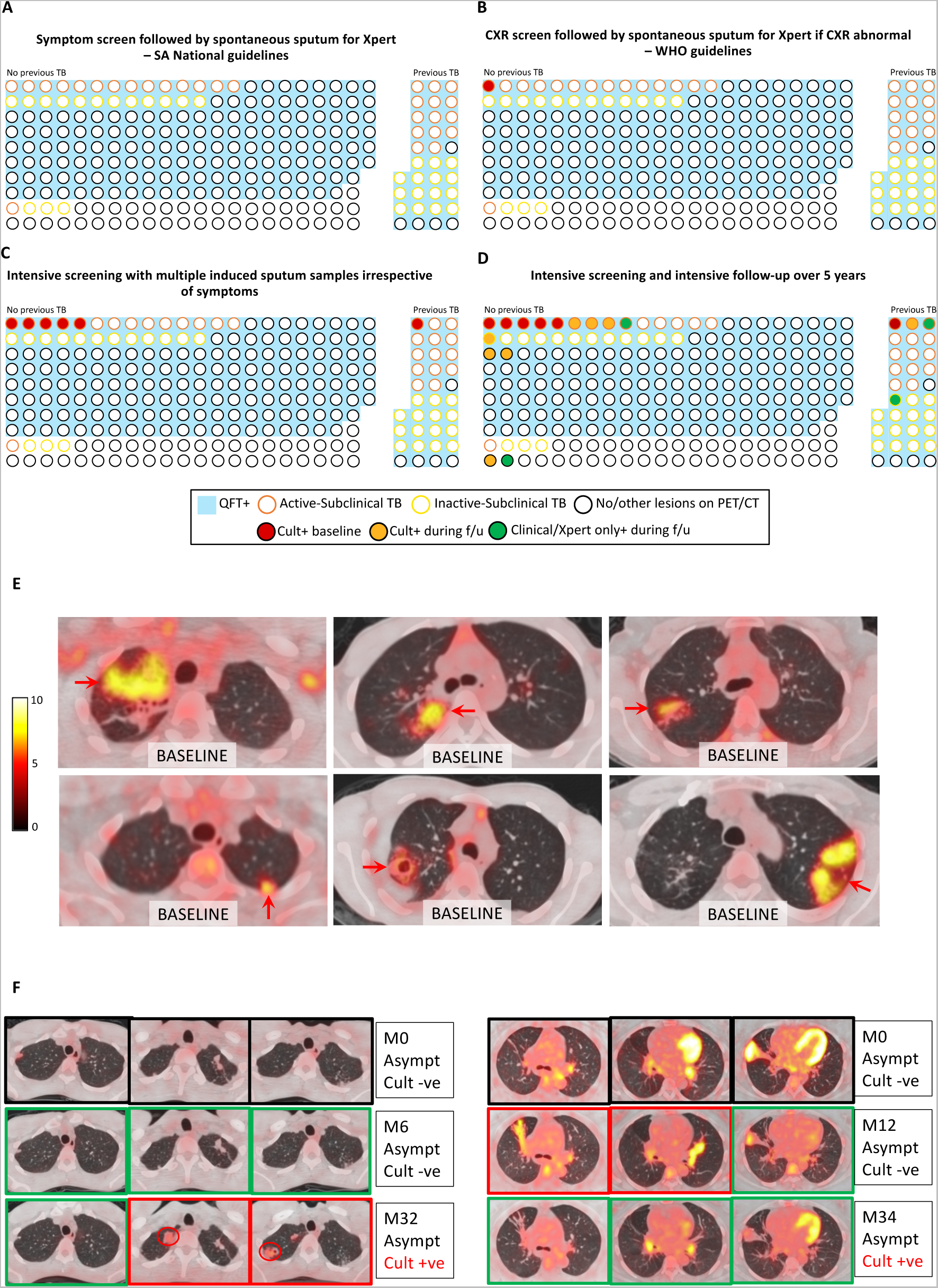
Difference in TB diagnosis according to different screening and follow-up approaches and PET/CT images of those diagnosed. Panels A to D show diagrammatic representation of the cohort by previous TB history and radiographic evidence of subclinical TB showing proportion with confirmed TB with different screening and follow-up approaches: (Panel A) South African guideline: Sputum investigation only if TB symptoms; (Panel B) WHO recommended approach using CXR with spontaneous sputum investigation only x-ray abnormal or symptoms; (Panel C) Intensive sputum investigation at baseline with 3x sputum (induced if needed); (Panel D) Intensive sputum investigation at baseline with follow-up over 5 years. Each of the 250 participants is represented by a circle with outline colour showing baseline PET/CT grouping (Subclinical – orange, Subclinical-inactive – yellow, no/other lesions – black). The circle is filled when TB is confirmed. The proportion with positive a Quantiferon test (QFT+) (i.e. with a clinical diagnosis of latent TB) and the proportion with previous TB is also represented. Cult+ = culture positive, f/u = follow-up. Panel E shows axial sections of fused FDG-PET/CT of the 6 culture positive participants at baseline. Panel F shows axial sections of fused FDG-PET/CT at baseline (top row), second PET/CT after 6-12 months (middle row) and at TB diagnosis (bottom row) of 2 participants finally diagnosed with TB at 32 (left) and 34 (right) months. Outline colour denotes; Black = baseline, Green = lesion improves or no change, Red = lesion worsens.

**Figure 3.**
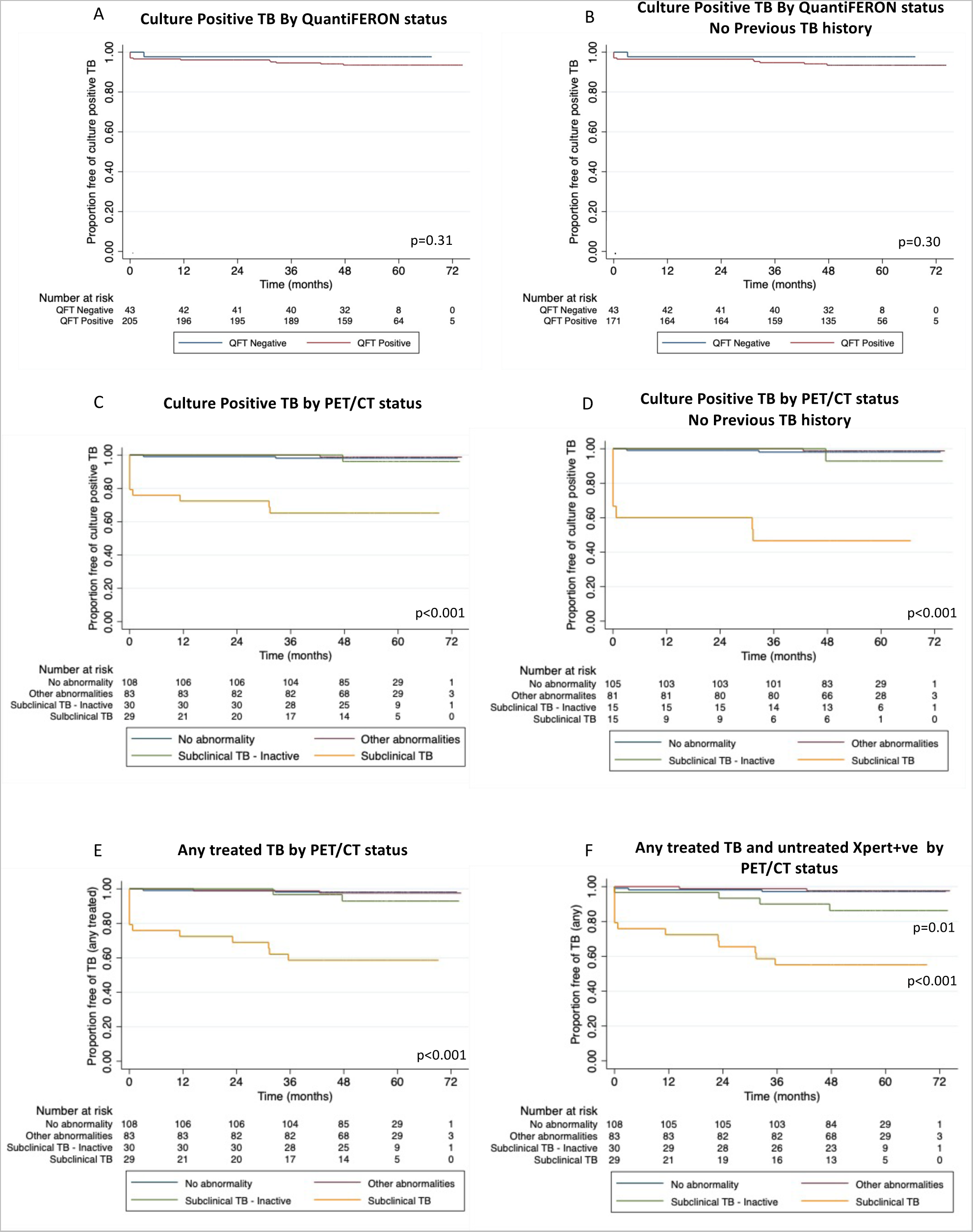
Survival curves for culture positive TB diagnosis according QuantiFERON or PET/CT findings. Survival curves showing development of TB over time by QuantiFERON status inducing (Panel A) or excluding (Panel B) those with no previous history of TB. Survival curves showing development of TB over time by nature of baseline radiography inducing (Panel C) or excluding (Panel D) those with no previous history of TB. Survival curves showing development of any treated TB (Panel E) or any treated TB and untreated Xpert-positive TB (Panel F) over time by nature of baseline radiography inducing those with no previous history of TB.

Of those with Subclinical TB on PET/CT 10/29 (34.5%) were ultimately diagnosed culture-positive (all when still asymptomatic), compared to 1/30 (3.3%) with Subclinical TB-inactive, 1/83 (1.2%) with other abnormalities and 2/108 (1.83%) with normal lungs (**Table 1**). The four culture-positive cases that did not have Subclinical TB were all drug-sensitive. Conversely, 7/10 culture-positive Subclinical TB cases had drug resistance similar to the index case; and three had index and contact isolates available with linkage confirmed by whole genome sequencing (**Supplementary Table S3**). The remaining 3/10 were drug sensitive. Comparison of the demographic and radiological features of those with Subclinical TB who did, and did not, develop culture positive TB is shown in **Supplementary Table S4**.

Compared to those with normal lung parenchyma, those with Subclinical TB on PET/CT (all of whom would not have been picked up by South African screening guidelines) were at significantly increased risk of culture positive TB, Hazard Ratio (HR) 22.37 (4.89-102.47), p<0.001. In a model adjusting for previous TB, the HR increased to 39.7 (8.42-187.33), p<0.001 (**Table 2 and Figure 3C-D**). Excluding one participant who would have been diagnosed with the WHO screening approach or six participants diagnosed culture positive by the study approach at baseline, Subclinical TB on baseline PET/CT remained significantly associated with development of culture positive TB during follow-up, HR 21.29 (4.59-98.80) p<0.001 and HR 10.65 (1.94-58.22) p=0.006, respectively. Expanding the TB case definition to include the four additional treated TB cases and then the four additional untreated TB cases had little impact on HR for Subclinical TB on PET/CT, HR 28.10 (6.27-125.85) and HR 20.65 (5.87-72.68), respectively (**Figure 3E-F and Supplementary Results-2.5**). By contrast those with positive QFT were not at significant increased risk of culture positive TB compared to those with a negative QFT HR 2.76 (0.36-21.13), p=0.33 (**Figure 3A-B and Supplementary Results-2.4)**.

**Table 2.** Univariate and multivariate analyses of the risk to develop culture positive TB over the study period due to baseline PET/CT parenchymal abnormalities and main covariates. N=247.

| Variable | Category | Univariate |  | Multivariate model adjusting for previous TB |  |
| --- | --- | --- | --- | --- | --- |
|  |  | Hazard Ratio (95% CI) | p value | Hazard Ratio (95% CI) | p value |
| Baseline PET/CT | Normal Lung | REF |  | REF |  |
|  | Subclinical TB | **22.37 (4.89-102.47)** | **<0.001** | **39.7 (8.42-187.33)** | **<0.001** |
|  | Subclinical TB-Inactive | 1.73 (0.16-19.08) | 0.65 | 2.83 (0.25-31.5) | 0.40 |
|  | Other lung abnormalities | 0.63 (0.06-6.90) | 0.70 | 0.62 (0.06-6.87) | 0.70 |
| Previous TB | No | REF |  | REF |  |
|  | Yes | 1.06 (0.24-4.75) | 0.94 | **0.19 (0.04-0.87)** | **0.03** |
| Age | Per year | 1.01 (0.97-1.05) | 0.77 |  |  |
| Sex | Female | REF |  |  |  |
|  | Male | 1.14 (0.39-3.27) | 0.81 |  |  |
| Hours of contact with index | ≤6hours/day | REF |  |  |  |
|  | >6 hours/day | 2.10 (0.73-6.05) | 0.17 |  |  |
| Any QuantiFERON | Negative | REF |  |  |  |
|  | Positive | 2.76 (0.36-21.13) | 0.33 |  |  |

### Performance of baseline PET/CT findings to predict progression from infection to disease

Thirteen of the 14 confirmed with culture-positive TB over the study period were clinically defined by QFT as having latent TB infection at baseline of which 10 (76.9%) had Subclinical TB on baseline PET/CT. In those with a clinical diagnosis of latent TB infection in whom TB disease was microbiologically excluded by the WHO screening approach, performance of PET/CT-defined Subclinical TB for culture positive TB over a 3-year period was; PPV 33.3% (95%CI 16.5%-54%), sensitivity 90% (95%CI 55.5%-99.7%) and specificity 90.7% (95% CI 85.7%-94.4%) exceeding the optimal WHO target product profile (TPP) characteristics for a test that predicts progression from TB infection to active disease.^19^

### Change in parenchymal lesions over time

112/250 participants had a repeat PET/CT after a median of 203 days (IQR 188-287), and of these 110 had not received any TB treatment prior to repeat PET/CT. The proportion of untreated participants showing a noticeable change in parenchymal lesions (**Supplementary Methods**) was significantly different in relation to the nature of lesions at baseline (p<0.001). Three quarters (12/16) of those with Subclinical TB showed substantial (greater than minimal) changes over time (6 worsening/mixed, 6 improving), compared to 23.5% (4/17) of those with Subclinical TB-inactive lesions (4 worsening/mixed) and 17.5% (7/40) with other lesions (5 improving, 2 worsening/mixed). By contrast there was no significant difference in noticeable changes in lymph nodes related to the nature of parenchymal lesions at baseline (p=0.136) **(Supplementary Figure S2).** Twenty-eight participants underwent bronchoalveolar lavage following the repeat PET/CT all of which were culture negative (**Supplementary Results-2.2**).

Eight of those who had a repeat PET/CT after a median of 248.5 days [IQR 192.5-371.0] were subsequently treated for TB (5 culture positive and 3 clinically diagnosed) a median 669 days [IQR 222-842]) after the repeat scan. Between baseline and second PET/CT, 2 showed improvement, 3 mixed, 3 minimal improvement/no change. Four had a third scan at the time of TB diagnosis, 2 showed improvement and 2 worsening, compared to second PET/CT (**Figure 2 and Supplementary Figure S1B-C**).

## Discussion

This study is the largest to systematically use high resolution imaging in asymptomatic contacts of TB and the first with prolonged follow-up in the absence of treatment and builds on our previous work to characterise and develop definitions for subclinical TB lesions.^15, 20^ We found that over three-quarters (76.9%) of asymptomatic, HIV-uninfected adults with a clinical diagnosis of latent TB who were confirmed to have culture positive pulmonary TB over a 5-year period had radiologically apparent infiltrative disease at baseline. Overall, of the 11.6% (29/250) who had baseline radiographic findings consistent with Subclinical TB disease, 34.5% (10/29) were subsequently diagnosed with culture positive TB, with 7/10 (70%) having drug resistance/WGS concordant with the index case. We have shown that slow evolution of radiographically evident disease pathology can occur over many years without apparent clinical manifestations.

Current tests for latent TB infection poorly predict the development TB disease resulting in high numbers needed to be treated to prevent a case of disease. The WHO have developed a TPP for diagnostic tests that could better predict TB progression.^19^ In those with a clinical diagnosis of latent TB in whom TB disease was excluded, subclinical changes on PET/CT had a sensitivity of 90% and specificity of 91% for culture positive TB over three years exceeding the TPP optimal performance characteristics. Whilst such performance has not previously been achieved, PET/CT is not a feasible routine diagnostic. However, our findings do support its use as a reference standard to identify and validate blood-based biomarkers to accelerate development of such a test.

In our study, TB diagnosis was frequently made by intensive active case detection and induced sputum collection. It is conjectural what the natural history of such cases would have subsequently been; some may have self-healed, whilst it is also plausible that others would have eventually presented symptomatically. However, by contexualising our results with different screening scenarios we demonstrate that asymptomatic culture confirmable cases are being missed routinely. Our results mirror findings from the macaque model of TB from which a subclinical “percolator” phenotype has previously been described.^21^ It is also possible that individuals with subclinical disease could intermittently shed bacilli in respiratory secretions thus posing a transmission risk; in a recent study by Williams *et al* five adults screened in South Africa with negative sputum culture and a positive facemask *Mtb* DNA sample all had abnormalities on PET/CT.^22^

There are limitations to our study; it was conducted in a high TB burden setting in HIV-uninfected, adult contacts of DR-TB and hence we can only speculate on how our findings relate to other populations. Disease trajectories in the context of immunosuppression (e.g. with HIV co-infection) differ, potentially with higher proportions of those with Subclinical TB progressing. In low-incidence settings the likelihood of participants having a previous history of TB and having additional TB exposure during follow-up would be low. In our study the four cases of culture-positive TB in those without Subclinical TB changes were drug-sensitive (i.e. unlikely related to index) with one case being QFT-negative at baseline and the remaining three occurring after 32 months suggesting that exposures after the baseline PET-CT may have contributed in these instances. Fourteen percent of our cohort had a previous history of TB. Residual changes of previously treated disease are difficult to distinguish from changes associated with new disease, however the risk of culture-confirmed disease in those with Subclinical TB was higher in younger individuals and those without previous TB history. For these reasons in a low incidence setting the performance of PET/CT may be improved.

Our results challenge present concepts of the evolution of human TB disease having implications for diagnostic and intervention strategies. Although 82% had a diagnosis of latent TB infection, the 11.6% with baseline Subclinical TB carried most of the 5-year disease risk.

Developing diagnostics and therapeutic approaches that target this population would therefore be likely to result in a reduction in the numbers needed to treat to prevent a case of clinical disease and enable TB care and prevention strategies to drastically limit onwards transmission and potential post-TB sequelae.

## Data Availability

All data produced in the present study are available upon reasonable request to the authors.

## Acknowledgements

Funding for this study was provided by the South African Medical Research Council (SAMRC; SHIP-02-2013), NIH (U19AI111276 and R01106804), Bill and Melinda Gates Foundation (37822), Wellcome (203135Z/16/Z) and WEHI (BDO innovation fund and philanthropy). RJW is supported the Francis Crick Institute which receives funding from Wellcome (CC2012), Cancer Research UK (CC2012) and MRC (FCC2012); AKC is supported by the NHMRC (GNT2020750); MB is supported by the NHMRC (GNT1195236); LEV and CEB were supported by the Division of Intramural Research, NIAID, NIH. The authors thank the Provincial Health Department of the Western Cape for use of the Site B health facility in Khayelitsha and for access to the Provincial Health Data Centre, with particular thanks to Mariette Smith and Nicki Tiffin. The authors thank Stephen Wilcox of the Advanced Genomics Facility at the Walter and Eliza Hall Institute of Medical Research for support and assistance in this work, which was made possible through Victorian State Government Operational Infrastructure Support and Australian Government NHMRC IRIISS. For the purposes of open access, the authors have applied a CC-BY public copyright to any author-accepted manuscript arising from this submission.

