## Supplementary materials for "High resolution imaging and five-year tuberculosis contact outcomes"

#### Supplementary Appendix

#### Table of contents

1. SUPPLEMENTARY METHODS 2

1.1. DEFINITION OF HOUSEHOLD CONTACTS (HHC) 2
1.2. DETAILED DESCRIPTION OF RECRUITMENT, CONSENT AND PROCEDURES 2
1.3. FOLLOW UP FOR ACTIVE TB 3
1.4. CHEST RADIOGRAPH READING 4
1.5. QUANTIFERON ASSAYS 4
1.6. FDG-PET/CT 5
1.6.1. PERFORMING FDG-PET/CT SCANS 5
1.6.2. READING FDG-PET/CT SCANS 5
1.6.3. RADIOGRAPHIC CATEGORIZATION OF PARTICIPANTS 6
1.6.4. CATEGORIZATION OF CHANGE BETWEEN BASELINE PET/CT SCAN AND FOLLOW-UP PET/CT SCAN 7
1.7. BRONCHOSCOPY 8
1.8. SPUTUM PROCESSING FOR MYCOBACTERIUM TUBERCULOSIS 8
1.9. MYCOBACTERIUM TUBERCULOSIS CULTURE FOR DNA EXTRACTION AND EXTENDED DRUG SENSITIVITY TESTING 9
1.10. MYCOBACTERIUM TUBERCULOSIS WHOLE GENOME SEQUENCING 10
1.11. MYCOBACTERIUM TUBERCULOSIS WGS DATA PROCESSING AND ANALYSIS. 10

2. SUPPLEMENTARY RESULTS 12

2.1. DRUG SUSCEPTIBILITY TESTING 12
2.2. BRONCHOALVEOLAR LAVAGE CULTURE 12
2.3. LYMPH NODE ABNORMALITIES 12
2.3.1. RELATIONSHIP OF BASELINE LYMPH NODE FDG-AVIDITY TO DEMOGRAPHICS AND INDEX EXPOSURE 12
2.4. INCIDENCE OF TB 13
2.5. SENSITIVITY ANALYSES 13
2.5.1. SENSITIVITY ANALYSIS 1 - CULTURE POSITIVE AND OTHER TREATED TB 13
2.5.2. SENSITIVITY ANALYSIS 2 - ANY TB (TREATED OR UNTREATED) 14

3. SUPPLEMENTARY FIGURES 15

Figure S1. Radiographic and microbiological findings at baseline and TB diagnosis 18
Figure S2. Change on follow-up scan undertaken after 5-15 months in relation baseline scan. 19
Figure S3. Lymph node abnormalities and their relationship of index case exposure in those *Mtb* sensitisation. 20

4. SUPPLEMENTARY TABLES 21

Table S1. Distribution of lesion types and spatial location (bronchopulmonary segment) by baseline PET/CT radiographic category 21
A All participants 21
Table S2. Baseline characteristics, clinical and radiographic findings, and TB outcomes of HHC who underwent PET/CT with no previous TB diagnosis. 22
Table S3. Microbiological and clinical characteristics of participants with culture positive TB compared to their index case. 24
Table S4. Characteristics of participants with radiographic evidence of Subclinical TB in relation to development of culture positive TB. 25
Table S5. Univariate and multivariate analyses of odds of having FDG-avid lymph nodes on baseline PET/CT by main covariates. n=247 unless stated. 26

5. REFERENCES 27

#### 1. Supplementary Methods

##### *1.1. Definition of Household contacts (HHC)*

HHC were defined as individuals sleeping in the same dwelling or room and/or providing themselves jointly with food or other essentials for living during the day with the index case for at least 7 days (continuous or non-continuous) during the 3 months prior to the index case being diagnosed with TB.

##### *1.2. Detailed description of recruitment, consent and procedures*

Index cases aged ≥15 years with at least rifampin resistant (RR) pulmonary TB confirmed by Xpert MTB/RIF or culture consented to a household visit and to contact household members. At that visit a detailed description of the household's setting and composition were taken. Listed individuals who met the definition of a household contact over the age of 18 years interested in participation were given an appointment at the study clinic for screening.

Following initial consent at the screening visit, all HHC underwent medical history and examination, screening for HIV with point of care testing followed by HIV ELISA if negative, screening for TB with symptom screen aligned with the South African National Guidelines (unexplained cough for 2 weeks, fever, night sweats, weight loss), physical examination for signs of pulmonary and extrapulmonary TB, digital posterior anterior (PA) chest radiography (CXR) and three sputum samples (the first spontaneously produced if able with the remainder induced using nebulised 3% saline to a total of three) all three being sent for smear, Xpert MTB/RIF and culture irrespective of symptoms or CXR findings. In addition, random capillary plasma glucose was taken to screen for diabetes mellitus and in women of child bearing potential a urine sample was taken for pregnancy testing.

Of those screened a subgroup of 250 further consented to undergo FDG-PET/CT subject to the following pre-specified inclusion/exclusion criteria

- Inclusion criteria - age ≥ 18 years, willing to undergo HIV counselling and testing (HCT)
- Exclusion criteria - HIV infection, on TB treatment at the time of screening, symptoms or signs of active TB or acute illness, age >65 years, smoker >30 pack years, diagnosis of malignancy, diagnosis of chronic lung infection other than TB (e.g., non-tuberculosis

mycobacteria [NTM], Fungal), diagnosis of chronic inflammatory condition associated with pulmonary pathology (e.g., Sarcoid, RA, Wegener's granulomatosis, bronchiectasis), inhaled or systemic steroid use within previous 2 weeks, breast feeding, pregnant, or planning pregnancy over next 3 months, unable to be followed up for 6 months, uncontrolled diabetes mellitus, or clinician discretion.

The 250 participants undergoing FDG-PET/CT scanning also provided consent for additional blood sampling for full blood count (FBC), C-Reactive Protein (CRP), Erythrocyte Sedimentation Rate (ESR), QuantiFERON Gold in tube (QFT-Gold), and QuantiFERON Gold Plus (QFT-Plus). Bloods for additional immunological investigations, urine and sputum were processed and stored.

Following further consent, 112 of the 250 participants had repeat FDG-PET/CT imaging performed at 5-15 months. All participants in order of recruitment were invited to undergo a second scan, until approximately 100 were complete. We then enriched the sample with those who were either QFT negative with normal baseline scans and those with evident abnormalities to ensure the cohort undergoing repeat scans was well distributed. All those undergoing repeat FDG-PET/CT imaging had a sputum sample (induced if needed) sent for smear, Xpert MTB/RIF and culture. In addition, blood was taken and processed similarly to the baseline visit.

##### *1.3. Follow up for active TB*

All participants were contacts of drug resistant TB and, in line with South African and WHO recommendations which reflect the limited evidence base, preventive therapy was not offered with participants instead monitored closely for the development of active TB with referral to specialist services upon concern. Follow up of the entire screened cohort was both active and passive. The 250 participants were contacted by phone or SMS approximately every 3 months and advised to attend the research clinic for investigation if they had symptoms or signs of TB. They were seen in person at the research clinic at 12 months and assessed clinically for signs and symptoms of TB with sputum investigations conducted if symptomatic. All participants screened at baseline, including those who did not undergo FDG-PET/CT were seen at the clinic between 23 to 38 months and again actively screened for TB with three sputum samples (induced if needed) sent for smear, Xpert MTB/RIF and culture (irrespective of symptom status). In addition, all participants consented to interrogation of their health service encounters via the Provincial Health Data Centre

(PHDC) and clinic medical records to ensure that all episodes of TB that were diagnosed and/or treated within the Western Cape Province were captured.[1] The search of the PHDC record took place on 21st May 2021 to capture all TB episodes from the start of the study to this date and to confirm previous episodes prior to study enrolment.

Any of the 250 participants undergoing baseline FDG-PET/CT scan who developed active TB disease during follow-up were invited for further assessment which included history, examination, CXR and repeat FDG-PET/CT provided this did not result in more than 2 scans occurring within a 12 month period in order to limit exposure to ionising radiation.

Participants who were diagnosed with TB through research investigations were referred to the local TB clinic with relevant results and the final decision to treat and drug regimen was determined by the statutory TB service.

##### *1.4. Chest radiograph reading*

CXR were performed using a digital X-Ray machines (Delft Oldeca DR or Phillips Essenta DR) and captured posterior-anteriorly in full inspiration with the participant standing. The Digital CXR images were viewed on 2 megapixel screens using the OsiriXMD version 11.0.4 (Pixemo, Bernex, Switzerland) software package and reported by a medically qualified investigator blinded to clinical and microbiological status. CXR were fully assessed for evidence of TB and then classified as consistent with active TB, inactive TB, abnormal but not consistent with TB, or normal.

##### *1.5. QuantiFERON assays*

The QFT-Gold and QFT-Plus assays (Qiagen, Valencia, CA) were conducted and scored in accordance with manufacturer's instructions, with the addition of performing an 8-point 1/2 dilution series, to improve standard curve accuracy. For samples where the antigen stimulated IFN-ɣ value was >10 IU/ml and could not be accurately extrapolated from the standard curve, we diluted plasma up to 1/100 until a quantifiable result was obtained.

##### *1.6. FDG-PET/CT*

###### *1.6.1. Performing FDG-PET/CT scans*

FDG-PET/CT scans were performed at three different sites, all within approximately 15 miles from the study site in Khayelitsha; the Cape PET-CT centre in Panorama Mediclinic using a Siemens Biograph PET/CT machine, the Western Cape Academic PET-CT centre at Tygerberg provincial hospital using a Phillips Gemini PET/CT machine or the Cape Universities Body Imaging Centre (CUBIC) using a Siemens Biograph PET/CT machine. Imaging protocols were similar (PET Parameters: 120 kV, 200 mA, 0.75 seconds rotation time, and a pitch of 0.438 with a collimation of 16x0.75 mm. CT Parameters: 110 KV/200 Ma, I31s, Strength 3). All repeat imaging was performed on the same machine as the initial scan.

Participants undergoing FDG-PET/CT fasted for six hours prior to the scan and were escorted to the PET/CT centre by a research worker. Point Of Care (POC) blood sugar was performed and if ≥11.1mmol/L the scan was rescheduled. 2.8MBq/kg of FDG was administered via a cannula. Sixty minutes after FDG administration the PET/CT scan was performed. CT was limited to the thorax (neck to liver) to reduce radiation exposure. Total effective radiation dose per scan was approximately 10mSv (varying with body weight and height). The second PET/CT was performed using a similar dose of FDG and injection to scan time as the initial scan.

###### *1.6.2. Reading FDG-PET/CT scans*

Two independent structured reports blind to clinical history and microbiological results focusing on the lung parenchyma plus mediastinal and hilar lymph nodes were made. Detailed reporting instructions were provided to all readers. One report was provided by a nuclear medicine physician/radiologist and the second by infectious diseases physician experienced in the reporting and analysis of research PET/CT scans for tuberculosis. Differences between the two structured reports were resolved by a 3rd reader also blind to clinical history and microbiological results. The nuclear medicine physician/radiologist in addition provided a full clinical report to comment on additional findings.

Parenchymal lesions were categorised as infiltrates, fibrotic scars, apical scarring, active nodules or discrete nodules according to prespecified definitions below:

- Infiltrate: Irregular, air space opacification which may include presence of clusters of micronodules, tree-in-bud appearance or denser consolidation.
- Fibrotic scar: Linear fibrotic or fibro-cystic abnormalities slightly distorting the surrounding lung tissue with no radiographic signs of activity which could be calcified with only minimal FDG uptake.
- Apical scarring - minimal subpleural scars typically less than 10 mm long at the apex of upper lobe was distinguished from fibrotic scars.
- Nodules: Near-spherical opacity within the lung parenchyma which may have well- or poorly-defined edges and less than 3 cm in diameter, though typically 3-10 mm in size,
- Other lesions: Any other abnormality found in the lung parenchyma or pleura was captured as "other". In particular bronchiectasis, ground glass opacification and pleural thickening was recorded.

The size and location of lesions (bronchopulmonary segment) were described. The lesions were also evaluated for radiographic signs of disease activity (e.g. cavitation, tree-in-bud appearance, poorly defined margin). The density of the lesion was assessed by maximum Hounsfield Units (HU). FDG uptake within the parenchyma lesions was quantified by maximal standardised uptake value ($SUV_{max}$) and Visual Score (VS) (VS = 0 – No visible uptake of FDG, VS = 1 – FDG uptake within lesion greater that background lung parenchyma but less than mediastinal blood pool, VS = 2 – FDG uptake within lesion greater than mediastinal blood pool but less that liver, VS = 3 – FDG uptake within lesion greater than liver). Parenchymal lesions were considered to have abnormal FDG uptake if VS was $\geq 1$. $SUV_{max}$ and VS of mediastinal and hilar lymph nodes was assessed and FDG uptake was considered abnormal if VS $\geq 2$. Mediastinal and hilar LN were also assessed for size (considered abnormal if the short axis width was greater than 1 cm) and for evidence of mineralization. Abnormal lymph nodes were placed into one of the following lymph node basins following convention of the International Association for Study of Lung Cancer (IASLC); right or left superior mediastinal (IASLC 2 – 4), aortic (IASLC 5, 6), subcarinal (IASLC 7), inferior mediastinal subcarinal (IASLC 8, 9) and right or left hilar (IASLC 10 –14).

##### *1.6.3. Radiographic categorization of participants*

Following the consensus reading process outlined above, the baseline FDG-PET/CT scans were classified into the prespecified, mutually exclusive categories below on the basis of the radiographic findings alone. These definitions were based on findings from our previously published work which in turn reflect findings from previous imaging and autopsy studies on findings in early TB disease.[2-4,a]

- Radiographically consistent with TB disease (Subclinical TB) - Presence of infiltrate in bronchopulmonary segment R1, R2, R3, R6, L1/2, L3 or L6 **OR** Presence of nodule with VS ≥1 in bronchopulmonary segment R1, R2, R3, R6, L1/2, L3 or L6
- Radiographically consistent with inactive TB (Subclinical TB-inactive)- Presence of fibrotic scar (**NOT** isolated apical scar) in bronchopulmonary segment R1, R2, R3, R6, L1/2, L3 or L6
- Other abnormalities of uncertain significance - Infiltrates, scars or nodules in other bronchopulmonary segments **OR** other parenchymal lesions (e.g. bronchiectasis)
- Normal lung parenchyma – No radiographically visible lesions within the lung parenchyma. This group may have mediastinal and/or hilar lymph node abnormalities.

Metabolically active (FDG-avid) Lymph Nodes were defined as lymph nodes of any size with VS ≥ 2 in the mediastinal, hilar or aortic location. Hot lymph nodes could be present in combination with any of the above parenchymal categories

##### *1.6.4. Categorization of change between baseline PET/CT scan and follow-up PET/CT scan*

The change between the baseline and follow-up scan was determined for parenchymal lesions and lymph node lesions separately and classified as follows: [1]

- **No change** – No change in visual score of any lesions between baseline and follow-up scan
- **Minimal change** – Single lesion change in visual score +/- 1 between baseline and follow-up scan

---

[a] In the previously published study (Esmail *et al* Nat Med 2016) we grouped together those with infiltrates, active nodules and fibrotic scars into a single Subclinical TB category. In this study which had a larger sample size we separated those with infiltrates and active nodules (Subclinical TB) from those with fibrotic scars only (Subclinical TB-inactive) into 2 subgroup to better understand the trajectory and prognosis of these pathologies.

- **Improvement** – Single lesion reduction in visual score of >1 **OR**
  considerable reduction in size or SUVmax (as determined by reader) if visual score = 3 **OR**
  >1 lesion reducing by visual score = 1 **OR**
  complete resolution of lesion between scans
- **Worsening** – Single lesion increasing in visual score of >1 **OR**
  considerable increase in size or SUVmax (as determined by reader) if visual score = 3 **OR**
  >1 lesion increasing by visual score = 1 **OR**
  new lesion(s) developing between scans
- **Mixed** – Some lesions worsening while others improving

##### *1.7. Bronchoscopy*

To confirm whether *Mtb* could be isolated more frequently from subclinical TB lesions in the lung, 12 individuals with parenchymal FDG-PET/CT abnormalities indicative of Subclinical TB, 6 individuals with other parenchymal lesions and 10 individuals with no parenchymal abnormalities underwent bilateral PET-CT lesion guided bronchoscopy, performed at Tygerberg Hospital. 150ml of saline was first instilled into the contralateral lobe to the region of interest, followed by 150ml instilled into the lobe with lesions of interest. Approximately 50-70ml bronchoalveolar lavage fluid (BALF) was obtained from each lobe, and 10 ml from each lobe was sent for MGIT culture in the TB Genomics Laboratory, Tygerberg Campus, Stellenbosch University. Cultures were incubated for 42 days before being classified as negative.

##### *1.8. Sputum processing for Mycobacterium tuberculosis*

All sputum samples were processed in the accredited laboratories of the South African National Health Laboratory Services (NHLS) where auramine sputum smear, Xpert MTB/RIF (Cepheid, Sunnydale, CA) and mycobacteria growth indicator tube (MGIT) liquid TB culture (BD Diagnostic Systems) were performed. Cultures were incubated for 42 days before being classified as negative.

##### 1.9. *Mycobacterium tuberculosis culture for DNA extraction and extended drug sensitivity testing*

Culture positive MGIT tubes were transported to the Biosafety Level 3 (BSL3) Laboratory of the Institue of Infectious Disease and Molecualar Medicine, University of Cape Town. MGIT cultures were pelleted and *Mtb* was resuspended in 2ml 7H9/25% glycerol and two 1 ml aliquots stored at -80°C. Stocks were prepared for DNA extraction and identification of rifamicin (RIF) resistant colonies as previously described[5]. Brifely, glycerol stocks were innoculated in 5 ml 7H9/ADC/0.05 % Tween 80 and incubated at 37°C, once reached confluent growth after approximately 7-10 days, 500 µl of broth culture was plated on two 120 mm 7H10/OADC plates and one 7H10/OADC plate containing 1 µg/ml RIF (7H10/RIF) for selection of resistant single colonies. Plates were incubated at 37°C for up to 8 weeks and growth monitored. *Mtb* which grew on plates without RIF were scraped into one 50ml falcon containing 3 ml sterile H2O, once confluent. *Mtb* was heat-killed at 80°C in a waterbath for 1 hour, followed by phenol/chloroform DNA extraction, as previously described[6]. Cultures which grew on 7H10/RIF plates were recorded as RIF resistant and individual *Mtb* colonies which grew on 7H10/RIF were picked and inoculated into 5 ml 7H9/ADC/0.05 % Tween 80 containing 1 µg/ml Rifampicin and incubated at 37°C. Once reached confluent growth, 500 µl of broth culture was plated on two 7H10/OADC plates and plates incubated until confluent growth. To make a stock of the 7H9 RIF culture, 500 µl of the 7H9 broth culture was added to 500 µl of 50% glycerol and stored at -80°C. Once plates were confluent with growth, plates were scraped and heat-killed and DNA extracted as above.

For extended drug sensitivity testing, glycerol stocks from the original MGIT bulk cultures and those made from the RIF resistant colonies grown in 7H9 were sent to the TB Genomics Laboratory, Tygerberg Campus, Stellenbosch University. Phenotypic drug sensitivity was performed using the Sensititre MYCOTB MIC test (Thermo Scientific) for 12 first- and second-line anti-TB drugs: amikacin, cyclosporin, ethambutol, ethionamide, isoniazid, kanamycin, moxifloxacin, ofloxacin, para-aminosalicyclic acid, rifabutin, rifampicin, and streptomycin. Pyrazinamide susceptibility testing was performed using the BACTEC MGIT 960 method (BD Diagnostic Systems).

##### *1.10. Mycobacterium tuberculosis whole genome sequencing*

Prior to library preparation, DNA samples were cleaned by adding 50 µl genomic DNA solution with 40 µl (0.8x) magnetic beads (NucleoMag®). Samples were incubated for 5 minutes at room temperature (RT) to bind the DNA and then washed with 200 µl 80% ethanol twice while on a magnetic stand. Cleaned genomic DNA was eluted in 15 µl of nuclease-free water (Ambion). One hundred ng of cleaned DNA was fragmented using Fragmentase (NEB) following manufacturer's instructions with some modifications. Briefly, to fragment the genomic DNA, 20 ul solution containing DNA, *Fragmentase*, 10x buffer and $MgCl_2$ were incubated at 37 °C for 30 mins on a PCR machine. Enzymes were deactivated at 65 °C for 30 minutes. Unbound excess reagents were removed by diluting the sample to 40 µl with nuclease-free water followed by a bead clean-up using 1.5x volume of magnetic beads (NucleoMag®) and two washes of 80% ethanol. Finally, bound fragmented DNAs were eluted in 65 µl of nuclease-free water. Sixty µl of fragmented genomic DNA (100 ng) was indexed for Illumina sequencing using the TruSeq DNA sample Prep Kit (Illumina) as per manufacturer's instruction. Library concentration was quantified by Qubit™ dsDNA Assay kit and Picogreen Assay (Thermo Fisher Scientific) and library size was determined using TapeStation (Agilent). Equimolar amounts of libraries were pooled and diluted to 750pM for 150-bp paired-end sequencing on a NextSeq2000 instrument using the P2 300- cycle kit v2 chemistry (Illumina) as per manufacturer's instructions. To produce the sequences the base calling and quality scoring was performed by the Real Time Analysis (v2.4.6) software. The FASTQ file generation and de-multiplexing for the samples was performed by the bcl2fastq conversion software (v2.15.0.4).

##### *1.11. Mycobacterium tuberculosis WGS data processing and analysis.*

Alignment and variant calling were performed according to the GATK4 Germline short variant discovery best-practices workflow with joint-calling[7] against the *Mycobacterium tuberculosis* H37Rv reference genome (NC_000962), with the omission of "base quality score recalibration" and "variant quality score recalibration" steps. SNP variant calls were initially QC filtered based on the following VCF annotations: MQ > 30, FS < 60, SOR < 3, MQRankSum > -12.5, ReadPosRankSum > -8. To calculate pairwise SNP distances, the SNP variant callset was further filtered by first removing all samples with > 5% missing variant calls or median depth < 10X, then by removing variants with > 5% missing calls or median depth less than 3 standard deviations below the mean median variant depth. *Mtb* sublineages, mixed infections and drug resistance were then detected by an in-development software package TBtypeR[8]. Briefly, TBtypeR examines variant allele frequencies at a panel of lineage specific variant sites compiled from at Napier et al.[9], Thawornwattana et al.[10], Coscolla et al.[11] and Shuaib et al.[12] to assign *Mtb* sublineage and mixture frequencies. Following this, variant allele frequencies at known drug resistances sites derived from The 2021 WHO catalogue of *Mycobacterium tuberculosis* complex mutations associated with drug resistance[13] examined to assign drug resistance predictions.

#### 2. Supplementary Results

##### *2.1. Drug susceptibility testing*

In total 7 culture positive cases (4 baseline and 3 follow-up) amongst HHC were confirmed drug resistant, all had Active-Subclinical TB on baseline PET/CT (**Table S3**). Two were detected by bulk culture and 5 following selective culture on 7H10/RIF to identify minority resistant isolates. Three had both index and contact strains available for WGS, confirming linkage, with < 10 SNP different. One of these baseline cases was initially diagnosed drug sensitive with the index-matching strain (5-8 SNP different) only identified following treatment default after 3 months, indicating possible low level mixed infection at baseline. A fourth contact was confirmed by bulk WGS to have mixed infection at baseline: 97% sublineage 4.1.2.1 and 3% sublineage 4.3.2. The low abundance 4.3.2 isolate was determined MDR following plating the bulk culture on 7H10/RIF and resistant colonies cultured for phenotypic and genotype confirmation.

##### *2.2. Bronchoalveolar lavage culture*

Following the second PET/CT, 12 with Subclinical TB or Subclinical-inactive TB who had remained sputum-culture negative and 16 with other or no parenchymal lesions underwent bi-lateral bronchoalveolar lavage; first sampling the uninfected lobe(s) and then the contralateral lobe with evident lesions, when present. All samples were culture negative. One of these individuals subsequently progressed to symptomatic disease after a further 14 months, and one was sputum Xpert-positive culture-negative after a further 11 months, when still asymptomatic. Both had worsening Subclinical TB lesions on second PET/CT prior to bronchoscopy. This demonstrates the insensitivity of convential microbiological approaches to confirm TB disease processes during asymptomatic states.

##### *2.3. Lymph node abnormalities*

###### *2.3.1. Relationship of baseline lymph node FDG-avidity to demographics and index exposure*

Fifty-three participants (21.2%) had FDG-avid mediastinal and/or hilar lymph nodes. The presence of FDG-avid lymph nodes was associated with older age, QuantiFERON-positivity, Subclinical TB parenchymal changes, and spending >18 hours/day with the index case. With multivariate logistic regression the following remained significantly associated with the presence of FDG-avid LN; age >30 years (adjusted odds ratio (aOR) 2.46 (1.16-5.18) p=0.018), >18 hours per day contact with index case (aOR 5.80 (1.91-17.79) p=0.002), Subclinical TB-inactive- (aOR 5.78 (1.94 – 17.25) p=0.002, Subclinical TB aOR 9.44 (3.19 – 26.94), p<0.001) **(Table S5 and Fig. S3)**.

##### *2.4. Incidence of TB*

The overall incidence of culture-positive TB in those with latent TB (QFT positive) was 1.42/100py (95%CI 0.83-2.45), compared to 11.69/100py (95%CI 6.29-21.72) in those with Subclinical TB, and 0.36/100py (95%CI 0.12-1.12) in those without Subclinical TB, the later comparable to the TB incidence in QFT-negative participants 0.54/100py (95%CI 0.08-3.86).

##### *2.5. Sensitivity analyses*

The primary per protocol analysis was in relation to culture positive TB. Four additional cases were treated for TB by the statutory health service who had negative or unavailable culture results were included in sensitivity analysis 1. Four additional cases were Xpert positive but culture negative with no symptoms and not treated by the statutory health service (with no progression) and are included in sensitivity analysis 2. Survival curves for both analyses are shown in **Fig.3E-F**)

###### *2.5.1. Sensitivity analysis 1 - Culture positive and other treated TB*

| **Variable** | **Category** | **Univariate** | | **Multivariate model adjusting for previous TB** | |
|---|---|---|---|---|---|
| | | **HR (95% CI)** | **p value** | **HR (95% CI)** | **p value** |
| **PET/CT** | No abnormalities | REF | | REF | |
| | Other abnormalities | 1.26 (0.18-8.95) | 0.82 | 1.26 (0.18-8.93) | 0.82 |
| | Subclinical TB-inactive | 3.50 (0.49-24.82) | 0.21 | 5.15 (0.71-37.22) | 0.11 |
| | **Subclinical TB** | **28.10 (6.27-125.85)** | **<0.001** | **44.23 (9.50-205.80)** | **<0.001** |
| **Previous TB** | No | REF | | REF | |
| | Yes | 1.84 (0.61-5.58) | 0.28 | 0.33 (0.10-1.05) | 0.06 |

18 cases; 14 culture positive + 4 additional treated symptomatic TB (2 clinically diagnosed and 2 Xpert positive – 1 culture neg and 1 culture unavailable)

##### *2.5.2. Sensitivity analysis 2 - Any TB (treated or untreated)*

| **Variable** | **Category** | **Univariate** | | **Multivariate model adjusting for previous TB** | |
|---|---|---|---|---|---|
| | | **HR (95% CI)** | **p value** | **HR (95% CI)** | **p value** |
| **PET/CT** | No abnormalities | REF | | REF | |
| | Other abnormalities | 0.84 (0.14-5.01) | 0.85 | 0.83 (0.14-4.99) | 0.84 |
| | **Subclinical TB--inactive** | **4.81 (1.08-21.47)** | **0.04** | **6.40 (1.40-29.37)** | **0.01** |
| | **Subclinical TB** | **20.27 (5.76-71.33)** | **<0.001** | **28.95 (7.81-107.34)** | **<0.001** |
| **Previous TBx** | No | REF | | REF | |
| | Yes | 2.47 (0.97-6.32) | 0.06 | 0.45 (0.17-1.23) | 0.12 |

22 cases; 14 culture positive + 4 additional treated symptomatic TB + 4 asymptomatic cases with positive Xpert (culture negative).

#### 3. Supplementary Figures

##### Figure S1

**A**

**Baseline culture positive**

**Participant details** | **Baseline PET/CT** | **Baseline CXR** | **Follow-up PET/CT**

| Male 40-44y, QFT+ | Asymptomatic, Culture positive, Baseline | | |
|---|---|---|---|
| Previous TB | 1, DS, 2008 Tx Default | | |
| Baseline CXR | Abnormal: Active TB | | |
| Sputum 0m | Cul spont: 1/1<br>Cul induced:2/2 | GX spont: 1/1<br>GX induced:2/2 | SMR spont:0/1<br>SMR induced: 1/2 |
| Resistance 0m | Cul: DS; RR colony: RIF | | GX: NA |
| Index Resistance | Cul: DS | | GX: RIF |

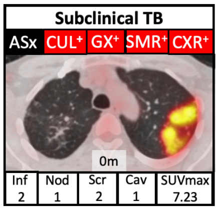

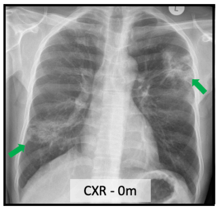

| Male 20-24y, QFT+ | Asymptomatic, Culture positive, Baseline | | |
|---|---|---|---|
| Previous TB | 0 | | |
| Baseline CXR | Abnormal: Active TB | | |
| Sputum 0m | Cul spont: 1/1<br>Cul induced:2/2 | GX spont: 0/1<br>GX induced:1/2 | SMR spont:0/1<br>SMR induced: 1/2 |
| Sputum 13m | Cul induced:1/1 | GX induced:1/1 | SMR induced: 1/1 |
| Resistance 0m | Cul: DS | | GX: DS |
| Resistance 13m | Cul: DS; RR colony: RIF PZA | | GX: DS |
| Index Resistance | Cul: RIF RBU | | GX: RIF |

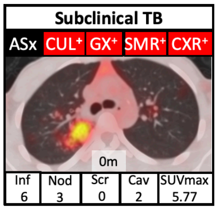

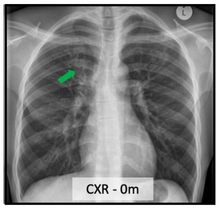

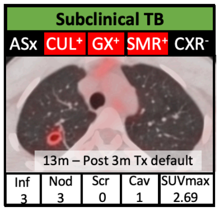

| Female 25-29y, QFT+ | Asymptomatic, Culture positive, Baseline | | |
|---|---|---|---|
| Previous TB | 0 | | |
| Baseline CXR | Abnormal: Active TB | | |
| Sputum 0m | Cul spont: 0/1<br>Cul induced:1/2 | GX spont: 0/1<br>GX induced:0/2 | SMR spont:0/1<br>SMR induced: 0/2 |
| Resistance | Cul: DS | | GX: NA |
| Index Resistance | Cul: RIF INH AMI(L) ETHAM(L) KAN(L) OFLX PZA | | GX: RIF |

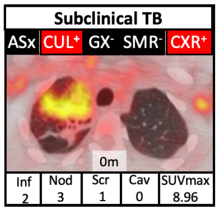

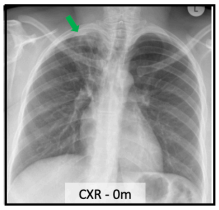

| Male 25-29y, QFT+ | Asymptomatic, Culture positive, Baseline | | |
|---|---|---|---|
| Previous TB | 0 | | |
| Baseline CXR | Abnormal: Active TB | | |
| Sputum 0m | Cul induced:1/3 | GX induced:0/3 | SMR induced: 0/3 |
| Resistance 0m | Cul: DS | | GX: NA |
| Index Resistance | Cul: RIF INH(L) ETHAM(L) PZA | | GX: RIF |

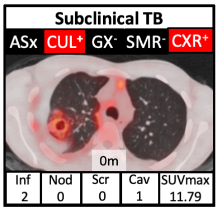

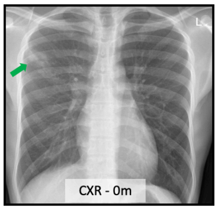

| Female 25-29y, QFT+ | Asymptomatic, Culture positive, Baseline | | |
|---|---|---|---|
| Previous TB | 0 | | |
| Baseline CXR | Abnormal: Active TB | | |
| Sputum 0m | Cul induced:2/3 | GX induced:0/3 | SMR induced: 0/3 |
| Resistance 0m | Cul: RIF INH PZA;<br>RR colony: RIF INH ETHAM(L) RBU PZA | | GX: NA |
| Index Resistance | Cul: RIF INH CYCLO ETHAM(L) RBU PZA | | GX: RIF |

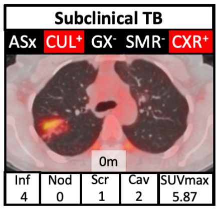

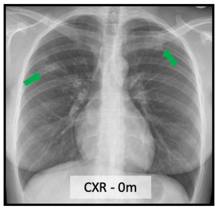

| Female 25-29y, QFT+ | Asymptomatic, Culture positive, Baseline | | |
|---|---|---|---|
| Previous TB | 0 | | |
| Baseline CXR | Normal | | |
| Sputum 0m | Cul induced:1/3 | GX induced:0/3 | SMR induced: 0/3 |
| Resistance 0m | Cul: DS;<br>RR colony: RIF RBU | | GX: NA |
| Index Resistance | Cul: RIF (RBU not tested) | | GX: RIF |

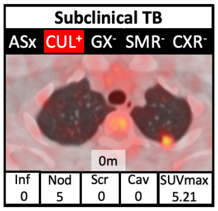

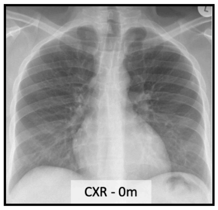

**B**

#### Follow-up culture positive

| Participant details |
|---|

Baseline PET/CT | Baseline CXR | PET/CT at diagnosis

| Male 55-59y, QFT+ | Asymptomatic, Culture positive, Month 1 | | |
|---|---|---|---|
| Previous TB | 0 | | |
| Baseline CXR | Abnormal: Active TB | | |
| Sputum 0m | Cul spont: 0/1<br>Cul induced:0/2 | GX spont:0/1<br>GX induced:0/2 | SMR spont:0/1<br>SMR induced: 0/2 |
| Sputum 1m | Cul induced: 1/1 | GX induced: 0/1 | SMR induced: 0/1 |
| Resistance 1m | Cul: DS;<br>RR colony: RIF INH RBU | | GX: NA |
| Index Resistance | Cul: RIF INH AMI OFLX | | GX: RIF |

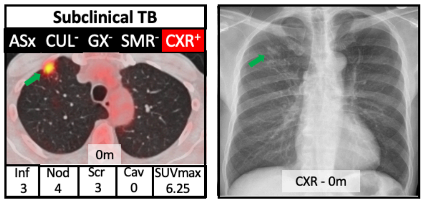

| Female 25-29y, QFT- | Symptomatic, Culture positive, Month 5 | | |
|---|---|---|---|
| Previous TB | 0 | | |
| Baseline CXR | Abnormal: Old TB | | |
| Sputum 0m | Cul spont: NA<br>Cul induced:0/3 | GX spont: NA<br>GX induced:0/3 | SMR spont: NA<br>SMR induced: 0/3 |
| Sputum 5m | Cul induced: 1/1 | GX induced: 1/1 | SMR induced: ND |
| Resistance 5m | Cul: DS | | GX: DS |
| Index Resistance | Cul: RIF INH STREP PZA | | GX: RIF |

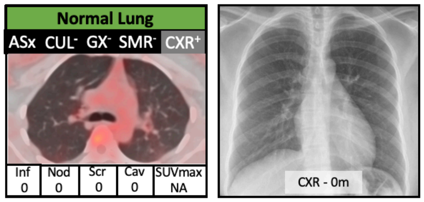

| Female 30-34y, QFT+ | Asymptomatic, Culture positive, Month 11 | | |
|---|---|---|---|
| Previous TB | 1: DS, 2009 Tx default | | |
| Baseline CXR | Normal | | |
| Sputum 0m | Cul spont: 0/1<br>Cul induced:0/2 | GX spont: 0/1<br>GX induced:0/2 | SMR spont:0/1<br>SMR induced: 0/2 |
| Sputum 11m | Cul induced: 1/1 | GX induced: 0/1 | SMR induced: 0/1 |
| Resistance 11m | Cul: DS | | GX: NA |
| Index Resistance | Cul: RIF INH | | GX: RIF |

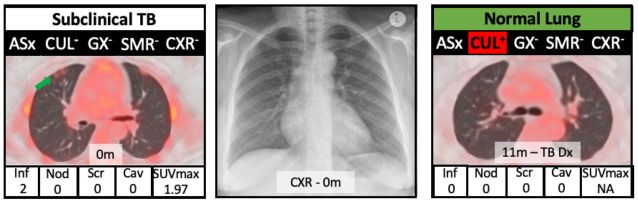

| Male 40-41y, QFT+ | Asymptomatic, Culture positive, Month 32 | | |
|---|---|---|---|
| Previous TB | 0 | | |
| Baseline CXR | Normal | | |
| Sputum 0m | Cul spont: 0/1<br>Cul induced:0/2 | GX spont: 0/1<br>GX induced:0/2 | SMR spont:0/1<br>SMR induced: 0/2 |
| Sputum 6m | Cul induced: 0/1 | GX induced: 0/1 | SMR induced: 0/1 |
| Sputum 32m | Cul spont: 1/1<br>Cul induced:3/3 | GX spont: 1/1<br>GX induced:1/3 | SMR spont: 0/1<br>SMR induced: 0/3 |
| Resistance 32m | Cul: RIF INH(L) STREP ETHIO | | GX: RIF |
| Index Resistance | Cul: RIF INH(L) STREP(L) PZA RBU | | GX: RIF |

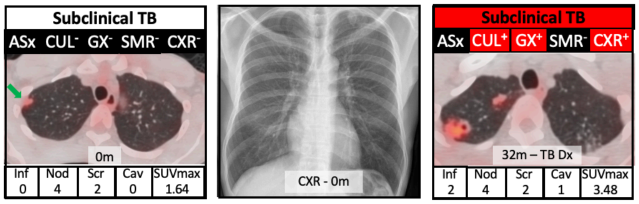

| Female 50-54y, QFT+ | Asymptomatic, Culture positive, Month 32 | | |
|---|---|---|---|
| Previous TB | 0 | | |
| Baseline CXR | Normal | | |
| Sputum 0m | Cul spont: 0/1<br>Cul induced:0/2 | GX spont: 0/1<br>GX induced:0/2 | SMR spont:0/1<br>SMR induced: 0/2 |
| Sputum 7m | Cul induced: 0/1 | GX induced: 0/1 | SMR induced: 0/1 |
| Sputum 32m | Cul spont: 0/1<br>Cul induced:1/2 | GX spont: 0/1<br>GX induced:0/2 | SMR spont: 0/1<br>SMR induced: 0/2 |
| Resistance 32m | Cul: DS | | GX: NA |
| Index Resistance | Cul: RIF INH STREP | | GX: RIF |

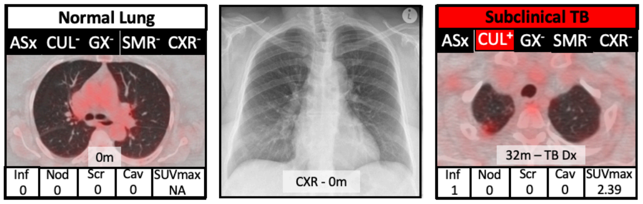

| Female 45-49y, QFT+ | Asymptomatic, Culture positive, Month 34 | | |
|---|---|---|---|
| Previous TB | 0 | | |
| Baseline CXR | Abnormal: Other | | |
| Sputum 0m | Cul spont: 0/1<br>Cul induced:0/2 | GX spont: 0/1<br>GX induced:0/2 | SMR spont:0/1<br>SMR induced: 0/2 |
| Sputum 12m | Cul induced: 0/1 | GX induced: 0/1 | SMR induced: 0/1 |
| Sputum 34m | Cul induced: 2/2 | GX induced: 1/2 | SMR induced: 0/2 |
| Resistance 34m | Cul: DS; RR colony = RIF | | GX: DS |
| Index Resistance | Cul: NA | | GX: RIF |

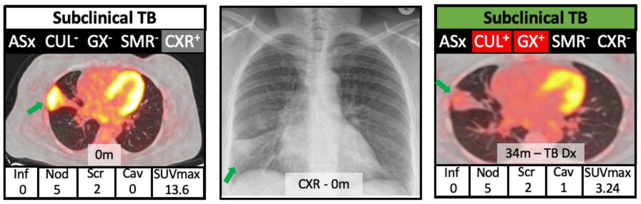

| Male 40-44y, QFT+ | Symptomatic, Culture positive, Month 48 | | |
|---|---|---|---|
| Previous TB | 0 | | |
| Baseline CXR | Abnormal: Old TB | | |
| Sputum 0m | Cul spont: 0/1<br>Cul induced:0/2 | GX spont: 0/1<br>GX induced:0/2 | SMR spont:0/1<br>SMR induced: 0/2 |
| Sputum 6m | Cul induced: 0/1 | GX induced: 0/1 | SMR induced: 0/1 |
| Sputum 48m | Cul spont: 1/2 | GX induced: 0/2 | SMR induced: 0/2 |
| Resistance | Cul: DS | | GX: NA |
| Index Resistance | Cul: RIF INH AMI OFLX | | GX: RIF |

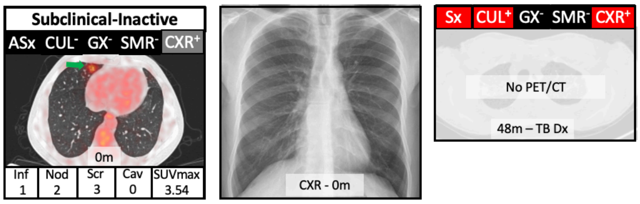

| Female 20-24y, QFT+ | Symptomatic, Culture positive, Month 53, HIV+ | | |
|---|---|---|---|
| Previous TB | 0 | | |
| Baseline CXR | Normal | | |
| Sputum 0m | Cul spont: 0/1<br>Cul induced:0/2 | GX spont: 0/1<br>GX induced:0/2 | SMR spont:0/1<br>SMR induced: 0/2 |
| Sputum 53m | Cul pleural Fluid:<br>1/1<br>Cul sputum: NA | GX pleural fluid: 0/1<br>GX sputum: 1/1 | SMR pleural fluid:<br>0/1<br>SMR sputum: 0/1 |
| Resistance | Cul: DS | | GX: DS |
| Index Resistance | Cul: RIF INH | | GX: RIF |

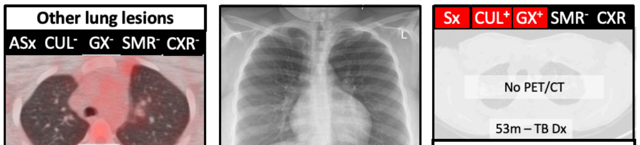

Pleural fluid Cul Pos
Sputum GX Pos (no cul)
Acquired HIV during fu

C

### Follow-up clinical diagnosis and treatment, culture negative

**Participant details** | **Baseline PET/CT** | **Baseline CXR** | **PET/CT at diagnosis**

| Female 25-29y, QFT- | Symptomatic, Xpert positive, Month 13 | | |
|---|---|---|---|
| Previous TB | 0 | | |
| Baseline CXR | Normal | | |
| Sputum 0 m | Cul spont: 0/1<br>Cul induced:0/2 | GX spont: 0/1<br>GX induced:0/2 | SMR spont:0/1<br>SMR induced: 0/2 |
| Sputum 13 m | Cul induced: 0/1 | GX induced: 0/1 | SMR induced: 0/1 |
| Sputum 15 m | Cul induced: NA | GX induced: 1/? | SMR induced: 0/? |
| Resistance 24 m | Cul: NA | | GX: RIF |
| Index Resistance | Cul: RIF INH | | GX: RIF |

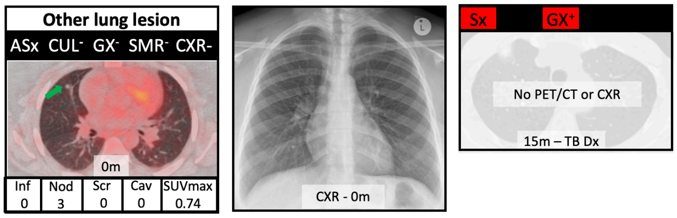

| Male 60-64y, QFT+ | Symptomatic, Clinical diagnosis, Month 24 | | |
|---|---|---|---|
| Previous TB | 1: DS, 2013 | | |
| Baseline CXR | Abnormal: Active TB | | |
| Sputum 0 m | Cul spont: 0/1<br>Cul induced:0/2 | GX spont: 0/1<br>GX induced:0/2 | SMR spont:0/1<br>SMR induced: 0/2 |
| Sputum 9 m | Cul induced: 0/1 | GX induced: 0/1 | SMR induced: 0/1 |
| BAL 11 m | RUL: 0/1 | LUL: 0/1 | |
| Sputum 16 m | Cul induced: 0/3 | GX induced: 0/3 | SMR induced: 0/3 |
| Sputum 24 m | Cul induced: 0/5 | GX induced: 0/5 | SMR induced: 0/5 |
| Resistance 24 m | Cul: NA – 6m DS treatment completed | | GX: NA |
| Index Resistance | Cul: RIF PZA | | GX: RIF |

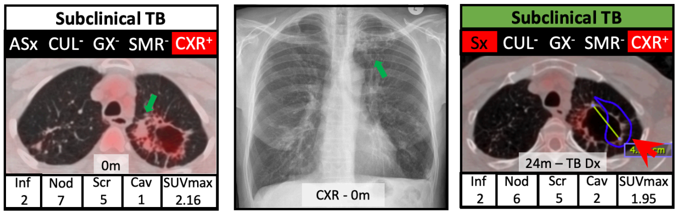

| Male 35-39y, QFT+ | Symptomatic, Xpert positive, Month 34, HIV+ | | |
|---|---|---|---|
| Previous TB | 1: 2006 | | |
| Baseline CXR | Abnormal: Active TB | | |
| Sputum 0m | Cul spont: 0/1<br>Cul induced:0/2 | GX spont: 0/1<br>GX induced:0/2 | SMR spont:0/1<br>SMR induced: 0/2 |
| Sputum 34m | Cul induced: 0/3 | GX induced: 1/3 | SMR induced: 0/3 |
| Resistance 34m | Cul: NA – 6m DS treatment completed | | GX: DS |
| Index Resistance | Cul: NA | | GX: RIF |

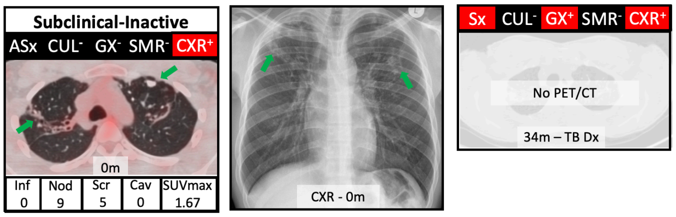

| Male 55-59y, QFT+ | Symptomatic, Clinical Diagnosis, Month 36 | | |
|---|---|---|---|
| Previous TB | 0 | | |
| Baseline CXR | Abnormal: Active TB | | |
| Sputum 0 m | Cul induced: 0/3 | GX induced: 0/3 | SMR induced: 0/3 |
| Sputum 6 m | Cul induced: 0/1 | GX induced: 0/1 | SMR induced: 0/1 |
| Sputum 24 m | Cul spont: 0/1<br>Cul induced:0/2 | GX spont: 0/1<br>GX induced:0/2 | SMR spont:0/1<br>SMR induced: 0/2 |
| Sputum 36 m | Cul : NA | GX : NA | SMR : NA |
| Resistance 36 m | Cul: NA, treated LEVO LZD BDQ, outcome unknown | | GX: NA |
| Index Resistance | Cul: RIF INH PZA RBU | | GX: RIF |

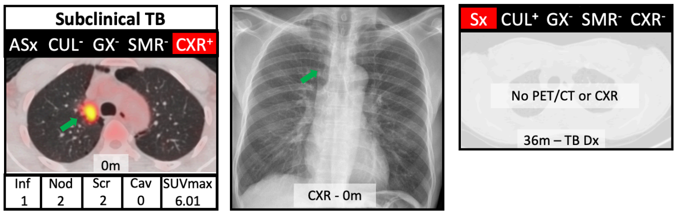

#### D Baseline and follow-up Xpert-positive culture-negative, no clinical diagnosis, no treatment.

**Participant details** | **Baseline PET/CT** | **Baseline CXR** | **PET/CT at diagnosis**

| Male 35-39y, QFT+ | Asymptomatic, Xpert positive, Month 0 | | |
|---|---|---|---|
| Previous TB | 1: DS, 2008 | | |
| Baseline CXR | Normal | | |
| Sputum 0m | Cul spont: 0/1<br>Cul induced:0/2 | GX spont: 1/1<br>GX induced:0/2 | SMR spont:0/1<br>SMR induced: 0/2 |
| Sputum 11m | Cul induced: 0/1 | GX induced: 0/1 | SMR induced: 0/1 |
| BAL 13m | Cul RUL: 0/1 | Cul LUL: 0/1 | |
| Resistance 0m | Cul: NA | | GX: DS |
| Index Resistance | Cul: DS | | GX: RIF |

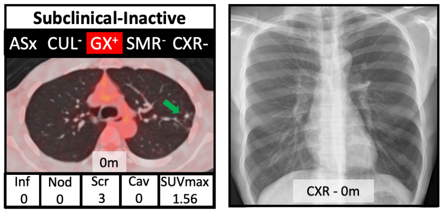

| Male 25-29y, QFT+ | Asymptomatic, Xpert positive, Month 0 | | |
|---|---|---|---|
| Previous TB | 0 | | |
| Baseline CXR | Normal | | |
| Sputum 0m | Cul spont: 0/1<br>Cul induced:0/2 | GX spont: 0/1<br>GX induced:1/2 | SMR spont:0/1<br>SMR induced: 0/2 |
| Sputum 11m | Cul induced: 0/1 | GX induced: 0/1 | SMR induced: 0/1 |
| Sputum 24m | Cul induced: 0/2 | GX induced: 0/2 | SMR induced: 0/2 |
| Resistance 0m | Cul: NA | | GX: DS |
| Index Resistance | Cul: DS | | GX: RIF |

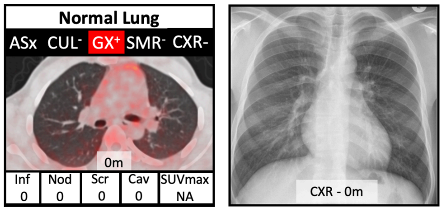

| Male 25-29y, QFT+ | Asymptomatic, Xpert positive, Month 24 | | |
|---|---|---|---|
| Previous TB | 2: DS, 2009; DS, 2013 | | |
| Baseline CXR | Abnormal: Active TB | | |
| Sputum 0m | Cul spont: 0/1<br>Cul induced:0/2 | GX spont: 0/1<br>GX induced:0/2 | SMR spont:0/1<br>SMR induced: 0/2 |
| Sputum 12m | Cul induced: 0/3 | GX induced: 1/3 | SMR induced: 0/3 |
| BAL 13m | Cul RLL: 0/1 | Cul LLL: 0/1 | |
| Sputum 24m | Cul spont: 0/1<br>Cul induced:0/2 | GX spont: 1/1<br>GX induced:0/2 | SMR spont:0/1<br>SMR induced: 0/2 |
| Resistance 24m | Cul: NA | | GX: DS |
| Index Resistance | Cul: RIF INH AMI OFLX | | GX: RIF |

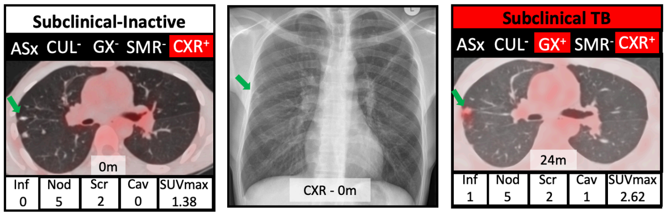

| Female 65-70y, QFT+ | Asymptomatic, Xpert positive, Month 23 | | |
|---|---|---|---|
| Previous TB | 0 | | |
| Baseline CXR | Normal | | |
| Sputum 0 m | Cul induced: 0/3 | GX induced: 0/3 | SMR induced: 0/3 |
| Sputum 10 m | Cul induced: 0/1 | GX induced: 0/1 | SMR induced: 0/1 |
| Sputum 24 m | Cul induced:0/3 | GX induced:1/3 | SMR induced: 0/3 |
| Resistance 36 m | Cul: NA | | GX: DS |
| Index Resistance | Cul: RIF INH FLUOROQUINOLONES SECOND LINE INJECTABLES | | GX: RIF |

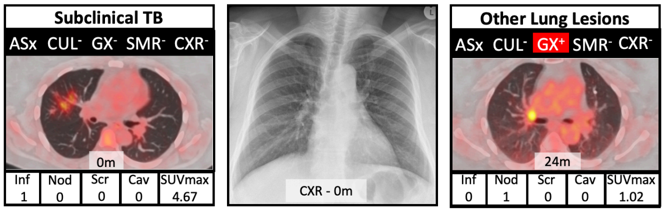

**Figure S1. Radiographic and microbiological findings at baseline and TB diagnosis**.

Shows details of all 22 participants diagnosed with TB by various definitions. For each, the table provides details of previous TB history, baseline chest X-ray (CXR) result and comprehensive microbiology of sputum and bronchoalveolar lavage (BAL, when performed), for each timepoint. The images show in the left column a single axial section from the baseline scan with (above image) detail of overall PET/CT category (top), details of symptoms (Sx, symptomatic; ASx, asymptomatic), positive (+) or negative (-) culture (CUL), Xpert (GX), and smear (SMR) and CXR result and (below image) number and nature of lesions (Inf, infiltrates; Nod, nodules; Scr, fibrotic scar; Cav, cavity; SUVmax, maximum parenchymal lesion standardized uptake value. The middle column shows the baseline CXR with lesions identified with green arrow. The right column shows single axial section at the point of diagnosis if occured during follow-up and available. The background colour of the overall PET/CT category on the top of this image indicates if the lesions were improved (green) or worsened (red) compared to the prior PET/CT.

**A** - baseline culture positive (n=6)

**B** - follow-up culture positive (n=8)

**C** - clinical diagnosis requiring treatment (n=4)

**D** - Xpert positive, culture negative, no clinical diagnosis and not treated (n=4).

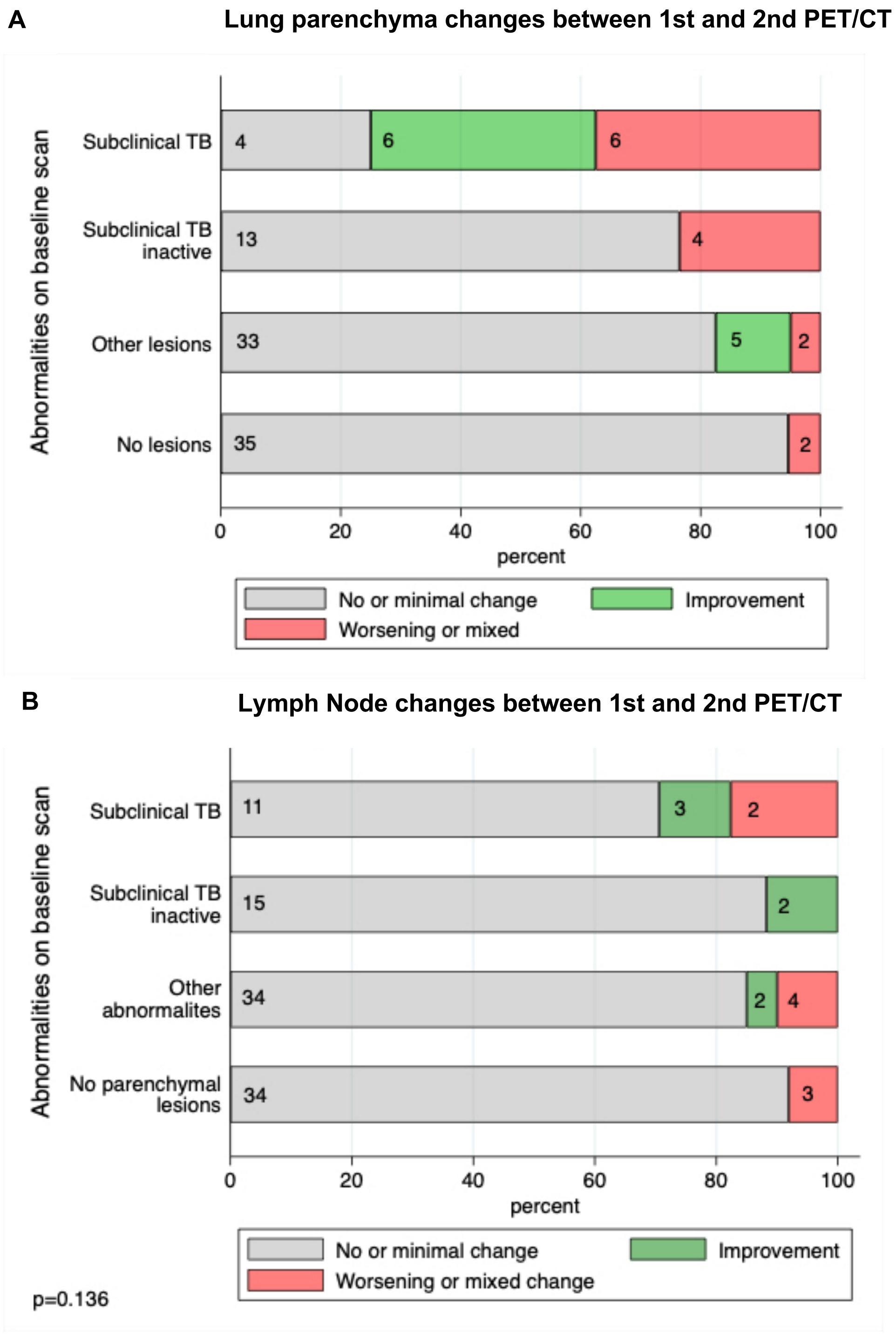

**Figure S2. Change on follow-up scan undertaken after 5-15 months in relation baseline scan.**

No or minimal change if there was a change in VS $\leq 1$ across all lesions. Improvement if there was a reduction in VS $\geq 2$ across all lesions (or clear improvement lesion with VS of 3) or resolution of a lesion. Worsening if there was an increase in VS $\geq 2$ across all lesions (or clear worsening in lesion with VS of 3) or development of a new lesion. Mixed if participant had individual lesions which showed a change in VS $\leq 1$ and other lesions with a change in VS $\geq 1$.

**A** - Parenchymal lesion changes at second PET/CT compared to first.

**B** - Lymph node lesion changes at second PET/CT compared to first.

A

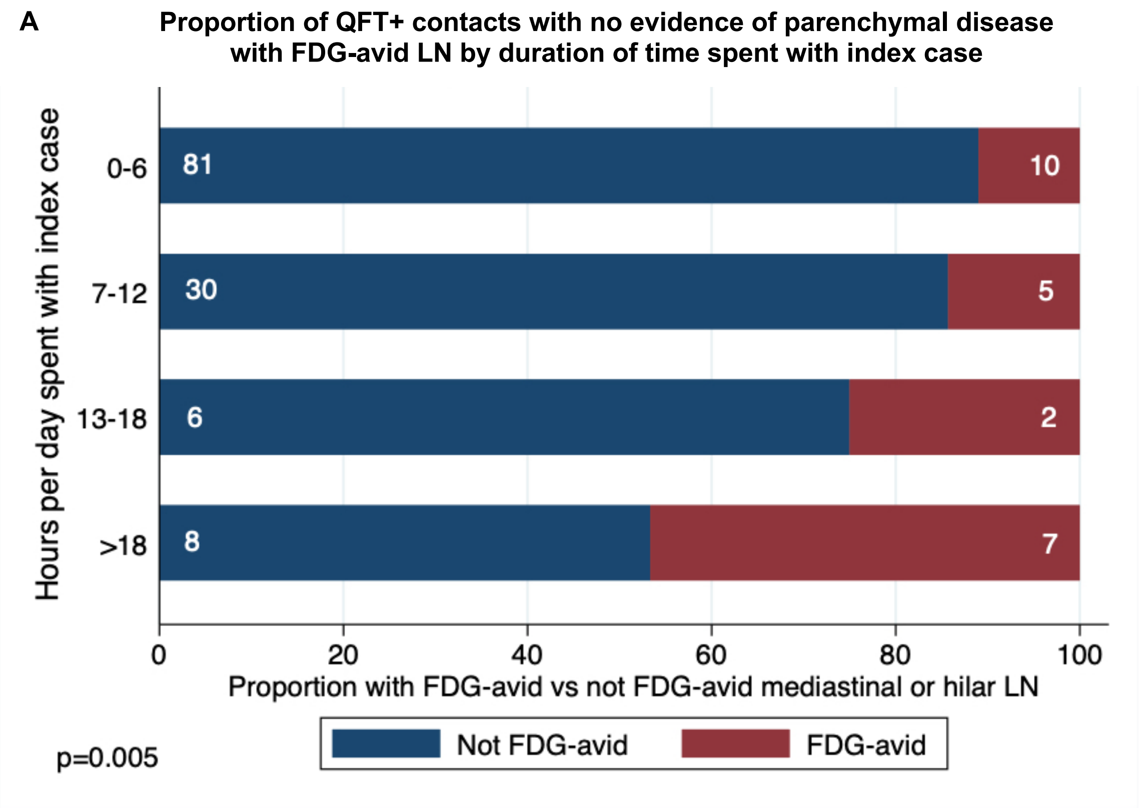

B

Example of lymph node lesions in those with no parenchymal lesions

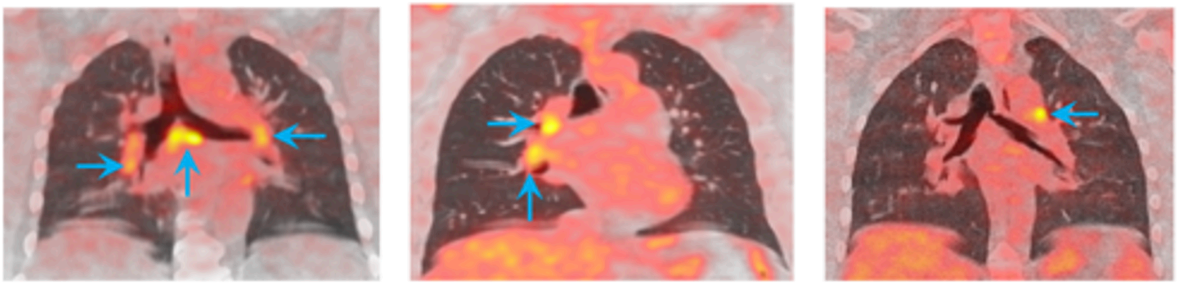

**Figure S3. Lymph node abnormalities and their relationship of index case exposure in those *Mtb* sensitisation.**

**A** - Proportion of QuantiFERON positive contacts with no FDG-PET/CT evidence of parenchymal disease but with FDG-avid lymph nodes (LN) by duration of time spent with index case.

**B** - Coronal sections of 3 participants with FDG avid lymph nodes with no subclinical parenchymal lesions (Left) subcarinal avid lymph nodes (Middle) hilar avid lymph nodes (Right) superior mediastinal avid lymph nodes**.**

#### 4. <u>Supplementary Tables</u>

##### Table S1. Distribution of lesion types and spatial location (bronchopulmonary segment) by baseline PET/CT radiographic category:

##### A All participants

| Lesion type | Total participants | Total lesions (av. number of lesions/ participant) | Infiltrates[%] (number with cavities) | Scars[%] (number with calcification) | Nodules[%] (number with calcification) | Bronchiectasis | Other | Right lung<br>RB1<br>RB2<br>RB3<br>RB6<br>RBOther | Left lung<br>LB12<br><br>LB3<br>LB6<br>LBOther |
|---|---|---|---|---|---|---|---|---|---|
| **Subclinical TB** | **29** | **183**<br>(6.31) | **45** [24.6%]<br>(8) | **43** [23.5%]<br>(13) | **89** [48.6%]<br>(26) | **4** | **3** | **23** [12.6%]<br>**23** [12.6%]<br>**24** [13.1%]<br>**5** [2.7%]<br>**27** [14.8%] | **35** [19.1]<br><br>**12** [6.6%]<br>**9** [4.9%]<br>**25** [13.7%] |
| **Subclinical TB-inactive** | **30** | **151**<br>(5.03) | **1** [0.7%]<br>(0) | **63** [41.7%]<br>(24) | **79** [53.2}<br>(45) | **5** | **3** | **24** [15.9%]<br>**11** [7.3%]<br>**17** [11.5%]<br>**8** [5.3%]<br>**17** [11.3%] | **29** [19.2%]<br><br>**19** [12.6%]<br>**4** [2.6%]<br>**22** [14.5%] |
| **Other** | **83** | **139**<br>(1.67) | **5** [3.6%]<br>(0) | **4** [2.9%]<br>(0) | **118** [87.8%] | **2** | **10** | **11** [7.9%]<br>**17** [12.2%]<br>**14** [10.1%]<br>**4** [2.9%]<br>**39** [28.1%] | **9** [6.5%]<br><br>**6** [4.3%]<br>**7** [5.0%]<br>**32** [23.0%] |
| **Normal** | **108** | **0**<br>(0) | **0** | **0** | **0** | **0** | **0** | **0** | **0** |
| **TOTAL** | **250** | **473**<br>(1.89) | **51** | **110** | **286** | **11** | **16** | | |

##### B Participants with no previous TB diagnosis

| Lesion type | Total participants | Total lesions (av. number of lesions/ participant) | Infiltrates[%] (number with cavities) | Scars[%] (number with calcification) | Nodules[%] (number with calcification) | Bronchiectasis | Other | Right lung<br>RB1<br>RB2<br>RB3<br>RB6<br>RBOther | Left lung<br>LB12<br><br>LB3<br>LB6<br>LBOther |
|---|---|---|---|---|---|---|---|---|---|
| **Subclinical TB** | **15** | **67**<br>(4.47) | **22** [32.8%]<br>(5) | **8** [11.9%]<br>(3) | **36** [53.7%]<br>(6) | **0** | **1** | **15** [22.4%]<br>**9** [13.4%]<br>**12** [17.9%]<br>**2** [3.0%]<br>**8** [11.9%] | **10** [14.9%]<br><br>**3** [4.5%]<br>**4** [6.0%]<br>**4** [6.0%] |
| **Subclinical TB-inactive** | **15** | **60**<br>(4) | **1** [0.7%]<br>(0) | **21** [35%]<br>(7) | **34** [53.2}<br>(20) | **3** | **1** | **10** [16.7%]<br>**4** [6.7%]<br>**7** [11.7%]<br>**3** [5.0%]<br>**7** [11.7%] | **12** [20.0%]<br><br>**3** [5.0%]<br>**1** [1.7%]<br>**13** [21.7%] |
| **Other** | **81** | **133**<br>(1.64) | **5** [3.6%]<br>(0) | **4** [3.0%]<br>(0) | **112** [84.2%]<br>(22) | **2** | **10** | **10** [7.5%]<br>**17** [12.8%]<br>**11** [8.3%]<br>**4** [3.0%]<br>**38** [28.6%] | **9** [6.8%]<br><br>**5** [3.8%]<br>**7** [5.3%]<br>**32** [24.0%] |
| **Normal** | **105** | **0**<br>(0) | **0** | **0** | **0** | **0** | **0** | **0** | **0** |
| **TOTAL** | **216** | **260**<br>(1.2) | **28** | **33** | **182** | **5** | **12** | | |

RB = Right bronchopulmonary segments 1-6 and other, LB = Left Bronchopulmonary segment 1-6 and other.

**Table S2. Baseline characteristics, clinical and radiographic findings, and TB outcomes of HHC who underwent PET/CT with no previous TB diagnosis.**

| | Variable | All n=216 | Subclinical TB n=15 | Subclinical TB-inactive n=15 | Other lung lesions n=81 | Normal lung n=105 | p= |
|---|---|---|---|---|---|---|---|
| DEMOGRAPHICS | Age (years) | 28 (23-42.5) | **43 (28-52)** [1,2] | **47 (25-56)** [1,2] | 29 (23-39) | 27 (22-40) | **0.003** |
| | Sex, Female n (%) | 133 (61.6%) | 8 (53.3%) | 11 (73.3%) | 51 (63.0%) | 63 (60.0%) | 0.69 |
| | Daily index contact, n (%) | 180 (83.3%) | 14 (93.3%) | 12 (80%) | 70 (86.4%) | 84 (80%) | 0.46 |
| | >6hrs/day with index, n (%) | 38.6% | 40% | 46.7% | 44.4% | 32.7% | 0.37 |
| | 1st degree relative, n (%) | 42.1% | 26.7% | 66.7% | 48.2% | 39.2% | 0.11 |
| | Smoking history, n (%) | 30.6% | 33.3% | 33.3% | 27.2% | 32.4% | 0.87 |
| | BMI | 28.53 (22.63-34.71) | 28.18 (19.57-32.44) | 29.03 (22.17-37.02) | 29.27 (22.37-33.49) | 28.36 (23.48-35.24) | 0.60 |
| INVESTIGATIONS | Proportion QFN+ Any , n (%) | 79.1% | 93.3% | 80.0% | 86.4% | 72.8% | 0.07 |
| | CXR - Suggestive of TB, n (%) | 18/215 (8.4%) | 7 (46.7%) | 1/14 (7.1%) | 2 (2.5%) | 8 (7.6%) | **<0.001** |
| | QFN Nil IFNγ (IU/ml) | 0.03 (0-0.13) | 0.08 (0.01-0.15) | 0 (0-0.08) | 0.03 (0-0.18) | 0.03 (0-0.12) | 0.26 |
| | QFN Ag-Nil (Gold) IFNγ (IU/ml) | 5.44 (0.5-27.45) | 6.98 (1.05-20.53) | 6.86 (0.92-19.44) | 7.63 (0.93-48.4) | 3.94 (0.13-19.17) | 0.11 |
| | QFN Ag-Nil (Plus-1) IFNγ (IU/ml) | 4.78 (0.33-26.45) | 6.89 (1.02-18.96) | 10.7 (0.98-22.92) | 5.53 (0.95-40.26) | 2.68 (0.13-16.57) | 0.13 |
| | QFN Ag-Nil (Plus-2) IFNγ (IU/ml) | 5.08 (0.44-29.86) | 12.61 (1.16-24.88) | 7.37 (0.77-20.15) | 6.49 (0.76-48.44) | 3.46 (0.23-20.5) | 0.13 |
| | CRP (mg/L) | 3 (1-6) | 4 (1.2-11) | 5 (3-8) | 2.3 (1-5) | 2 (1-6) | 0.05 |
| | CRP>=10mg/L, n (%) | 15.0% | 26.7% | 20.0% | 13.8% | 13.6% | 0.54 |
| | ESR (mm/Hr) | 14 (5-29) | **20 (8-33)** [1] | **27 (20-45)** [1,2] | 10.5 (4-30) | 13 (3-25) | **0.002** |
| | WCC ($x10^9$/L) | 5.9 (4.73-7.54) | 7.56 (6.68-8.32) | 5.33 (4.95-7.18) | 5.78 (4.53-7.37) | 5.79 (4.74-7.54) | 0.06 |
| | Neutrophils ($x10^9$/L) | 3.17 (2.21-4.54) | 4.8 (3.69-5.63) | 2.6 (2.1-4.48) | 2.94 (2.17-4.6) | 3.16 (2.25-4.36) | 0.05 |
| | N:L | 1.55 (1.1-2.22) | 2.54 (1.48-3.2) | 1.46 (1.1-2.37) | 1.5 (1.09-2.21) | 1.59 (1.07-2.04) | 0.07 |
| | N:M | 8.0 (6.22-10.38) | 10.62 (7.24-12.31) | 7.21 (5.68-8.97) | 8.02 (6.44-10.33) | 7.97 (6.13-9.79) | 0.12 |
| | L:M | 5.11 (4.05-6.26) | 4.05 (3.47-5.74) | 5.63 (3.44-6.55) | 4.99 (4.31-6.73) | 5.17 (4.06-6.26) | 0.20 |
| PET/CT IMAGING FINDINGS | Lung total parenchymal lesions | 1 (0-2) | **5 (2-6)** [2] | 3 (1-7) | 1 (1-2) | NA | **0.0001** |
| | Lung largest lesion size (mm) | 6.69 (4.44-21) | **36.9 (12.9-56.3)** [2] | **30 (19.8-40.6)** [2] | 5.27 (4-9.84) | NA | **0.0001** |
| | total infiltrates | 0 (0-0) | **1 (1-2)** [2,3] | 0 (0-0) | 0 (0-0) | NA | **<0.0001** |
| | total fibrotic scars | 0 (0-1) | **0 (0-2)** [2,3] | **1 (1-2)** [2] | 0 (0-0) | NA | **<0.0001** |
| | total nodules | 1 (0-3) | 3 (0-4) | 1 (0-3) | 1 (1-2) | NA | 0.31 |
| | total cavities (range) | 0 (0-2) | **0 (0-2)** [2,3] | 0 (0-0) | 0 (0-0) | NA | **<0.0001** |
| | Lung maximum VS | 0 (0-1) | **3 (1-3)** [2,3] | 0 (0-1) | 0 (0-0) | NA | **0.0001** |
| | Lung SUVmax | 1.23 (0.83-1.69) | **5.21 (1.64-6.25)** [2,3] | **1.69 (1.28-1.96)** [2] | 1.08 (0.8-1.36) | NA | **0.0001** |
| | Lung HUmax | 48 (-56-273) | **120 (41-481)** [2] | **555 (36-1190)** [2] | 13 (-134.5-181.5) | NA | **0.003** |
| | LN total lesions | 0 (0-2) | **2 (2-3)** [1,2] | **2 (0-5)** [1,2] | 0 (0-2) | 0 (0-1) | **<0.0001** |
| | Proportion with FDG-avid LN (%) | 19.4% | **66.7%** | **53.3%** | 18.5% | 8.6% | **<0.001** |
| | Largest abnormal LN (mm) | 8.4 (5.42-11) | **12 (10.2-14.9)** [1,2,3] | 8 (5.4-10.6) | 8.2 (5-10.3) | 7.8 (5.1-9.8) | **0.01** |
| | LN maximum VS | 1 (1-3) | **3 (2.5-3)** [1,2] | **2 (1-3)** [1,2] | 1 (1-3) | 1 (1-2) | **0.02** |
| | LN SUVmax | 2.5 (1.9-4.16) | **4.68 (3.24-6.36)** [1,2] | **2.96 (2.22-5.07)** [1] | 2.75 (1.6-3.47) | 2.11 (1.84-2.8) | **0.005** |
| | LN HUmax | 612.50 (157-1096.50) | 391 (168-1106) | 797 (158-1244) | 488 (136-1013) | 669.5 (252-1159) | 0.65 |
| TB OUTCOMES | Culture positive at baseline (treated) | 5 (2.3%) | **5 (33.3%)** | 0 (0%) | 0 (0%) | 0 (0%) | **<0.0001** |
| | Culture positive at follow-up (treated) | 7/215 (3.2%) | **3 (20%)** | 1 (6.7%) | 1 (1.2%) | 2/104 (1.9%) | **0.0014** |
| | Clinical TB at follow-up (treated) | 2/215 (0.93%) | 1 (6.7%) | 0 (0%) | 1 (1.2%) | 0/104 (0%) | 0.088 |
| | Xpert positive culture negative (not treated) | 2 (0.93%) | 1[5] (6.7%) | 0 (0%) | 0 (0%) | 1[4] (0.95%) | 0.099 |
| | Any TB or Mtb detected over study period | 16 (7.4%) | **10 (66.7%)** | 1 (6.7%) | 2 (2.5%) | 3 (2.9%) | **<0.0001** |
| | Symptoms at Any treated TB diagnosis | 5/14 (35.7%) | **1/9 (11.1%)** | 1/1 (100%) | 2/2 (100%) | 1/2 (50%) | **0.047** |
| | Symptoms at culture positive TB diagnosis | 3/12 (25%) | **0/8 (0%)** | 1/1 (100%) | 1/1 (100%) | 1/2 (50%) | **0.025** |

Values are n (%) or median (IQR). Denominator values are indicated in the column descriptor, or indicated in a cell when values were missing. Relationship between PET/CT categories and baseline characteristics analysed for categorical variables by $\chi^2$ or Fisher's exact test, numerical variables by Kruskal Wallis with *post hoc* analysis using Dunn's multiple comparison testing. Bold and superscript number indicates which *post hoc* numerical comparisons are significantly different (p<0.05): 1 = in comparison with no lung lesions, 2 = in comparison with other lung lesions,

3 = in comparison with Inactive-Subclinical TB. BMI, body mass index; Clinical TB, TB symptom positive *Mtb* culture and Xpert negative; CRP, C-reactive protein; CXR, chest X-ray; ESR, erythrocyte sedimentation rate; $HU_{max}$, maximum Hounsfield units; IFNγ, interferon-gamma; LN, lymph node; N:L, neutrophil:lymphocyte ratio in blood; N:M, neutrophil:monocyte ratio in blood; QFN+, QuantiFERON positive; $SUV_{max}$, maximum standardised uptake value; VS, visual score; WCC, whole cell count; Xpert, GeneXpert version 3.0: superscript 4 = detected at baseline, 5 = detected at follow-up.

**Table S3. Microbiological and clinical characteristics of participants with culture positive TB compared to their index case.**

| Baseline PET/CT | Previous TB Episodes | Months to TB | Sx at TB | Index Extended DST | HHC Extended DST | HHC SNP distance to index | Linkage |
|---|---|---|---|---|---|---|---|
| Subclinical TB | 0 | 0 | ASx | **RIF AMI(L) ETHAM(L) INH KAN(L) OFLX PZA** | DS (bulk) | NA | DST not linked (WGS NA) |
| Subclinical TB | 0 | 0 | ASx | **RIF RBU** | DS (bulk) at baseline diagnosis; **3m default then RIF PZA (RR colony)** | 740; 5-8 | Baseline not linked **Post-default linked WGS SNPs + DR** |
| Subclinical TB | 0 | 0 | ASx | **RIF INH CYCLO ETHAM(L) RBU PZA** | **RIF INH PZA (bulk) RIF INH ETHAM(L) RBU PZA (RR colony)** | 1-3 | **WGS SNPs + MDR linked** |
| Subclinical TB | 0 | 0 | ASx | **RIF** | DS (bulk) **RIF RBU (RR colony)** | NA | **DST linked** (WGS NA) |
| Subclinical TB | 0 | 0 | ASx | **RIF INH(L) ETHAM(L) PZA** | DS (bulk) | NA | DST not linked (WGS NA) |
| Subclinical TB | 1: 2008 (DS) default | 0 | ASx | **RIF (Xpert), DS (culture)** | DS (bulk) **RIF (RR colony)** | NA | **DST linked** (no index WGS) |
| Subclinical TB | 0 | 1 | ASx | **RIF INH AMI OFLX** | DS (bulk), mixed WGS, 97% 4.1.2.1, 3% 4.3.2 **RIF INH RBU (RR colony), sub-lineage 4.3.2** | NA | **RR colony DST linked** (no index WGS) |
| Subclinical TB | 1: 2009 (DS) default | 11 | ASx | **RIF INH** | DS (bulk) | NA | DST not linked (no index WGS) |
| Subclinical TB | 0 | 32 | ASx | **RIF INH(L) STREP(L) PZA RBU** | **RIF INH(L) STREP ETHIO (bulk)** | 2-3 | **WGS SNPs + MDR linked** |
| Subclinical TB | 0 | 34 | ASx | **RIF (Xpert)** | DS (bulk) **RIF (RR colony)** | NA | **DST linked** (no index WGS) |
| Subclinical TB-inactive | 0 | 48 | Sx | **RIF INH AMI OFLX** | DS (bulk) | NA | DST not linked (WGS NA) |
| Normal lung | 0 | 5 | Sx | **RIF INH STREP PZA** | DS (bulk) | NA | DST not linked (WGS NA) |
| Normal lung | 0 | 32 | ASx | **RIF INH STREP** | DS (bulk) | 780 | WGS SNPs + DS not linked |
| Other lung lesion | 0 | 53 | Sx | **RIF INH** | DS (bulk) | NA | DST not linked (WGS NA) |

ASx, asymptomatic; bulk, bulk culture; DS, drug sensitive; DST, drug sensitivity testing; DR drug resistant; HHC, household contact; Index, TB index case of contact; L, low level resistant; Linkage, whether index case and HHC are linked via DST or whole genome sequence (WGS) via single nucleotide polymorphism (SNP) distance (<10 SNPs) or DR or MDR (multi-drug resistant) SNP pattern, red indicates linked; NA, not avaliable; RR, rifampicin resistant; Sx, symptomatic. AMI, amikacin; CAP, capreomycin; CYCLO, cyclosporin; ETHAM, ethambutol; ETHIO, ethionamide; INH, isoniazid; KAN, kanamycin; LEV, levofloxacin; MXF, moxifloxacin; OLFX, ofloxacin; PZA, pyrazinamide; RBU, rifabutin; RIF, rifampicin; STREP, streptomycin.

**Table S4. Characteristics of participants with radiographic evidence of Subclinical TB in relation to development of culture positive TB.**

| | Variable | Subclinical TB diagnosed culture+ TB n=10 | Subclinical TB Never culture+¶ n=19 | p= |
|---|---|---|---|---|
| DEMOGRAPHICS | Age (years) | **31.5 (28-40)** | 49 (38-52) | **0.01** |
| | Sex, Female n (%) | 5 (50%) | 9 (47.4%) | 0.89 |
| | Previous TB history | **2 (20%)** | 12 (63.2%) | **0.05** |
| | Daily index contact, n (%) | 8 (80%) | 16 (84.2%) | 0.78 |
| | >6hrs/day with index, n (%) | 2.5 (2-3) | 2 (2-3) | 0.99 |
| | Smoking history, n (%) | 4 (40%) | 8 (42.1%) | 0.91 |
| | BMI | 22.88 (19.63-29.32) | 24.08 (21.38-29.52) | 0.64 |
| BLOOD RESULTS | Proportion QFN+ Any, n (%) | 10 (100%) | 18 (94.7%) | 0.46 |
| | CXR – Suggestive of TB, n (%) | 6 (60%) | 10 (52.6%) | 0.30 |
| | QFN Nil value | 0.08 (0.01-0.19) | 0.05 (0-0.12) | 0.73 |
| | QFN Ag-Nil (Gold) | 6.97 (3.11-16.36) | 19.49 (1.05-66.96) | 0.25 |
| | QFN Ag-Nil (Plus-1) | 6.64 (2.39-16.76) | 16.19 (1.02-52) | 0.38 |
| | QFN Ag-Nil (Plus-2) | 11.45 (2.58-21.42) | 20.15 (1.16-66.62) | 0.60 |
| | CRP (mg/L) | 5.5 (3-20) | 2 (1-11) | 0.09 |
| | CRP>=10 mg/L, n (%) | 3 (30%) | 5 (26.3%) | 0.83 |
| | ESR (mm/Hr) | **35 (20-45)** | 15 (7-29) | **0.03** |
| | WCC ($x10^9/L$) | **7.82 (7.26-8.32)** | 6.62 (5.11-7.83) | **0.003** |
| | Neutrophils ($x10^9/L$) | **5.09 (4.15-5.63)** | 3.89 (2.99-4.43) | **0.01** |
| | N:L | 2.39 (1.94-3.06) | 1.96 (1.29-2.45) | 0.10 |
| | N:M | 10.63 (5.72-12.55) | 8.4 (7.20-11.16) | 0.6 |
| | L:M | 3.54 (2.39-5.72) | 4.75 (3.64-6.17) | 0.09 |
| IMAGING FINDINGS | Lung total parenchymal lesions | 5.5 (5-8) | 6 (3-10) | 0.58 |
| | Largest lesion size (mm) | 43.15 (19.7-57.8) | 44.19 (28-56.3) | 0.97 |
| | total infiltrates | 2 (0-3.25) | 1 (0-2) | 0.25 |
| | total fibrotic scars | 1 (0-2) | 2 (0-3) | 0.33 |
| | total nodules | 3 (0-4.25) | 3 (1-6) | 0.48 |
| | total cavities | 0 (0-1.25) | 0 (0-0) | 0.10 |
| | Lung maximum VS | **3 (3-3)** | 2 (1-3) | **0.02** |
| | Lung SUVmax | **6.06 (5.21-8.96)** | 2.16 (1.72-3.6) | **0.006** |
| | Lung HUmax | 152.5 (85-481) | 205 (88-782) | 0.54 |
| | LN total lesions | 1.5 (0-3.25) | 2 (0-3) | 0.91 |
| | Proportion with FDG-avid LN, n (%) | 6 (60%) | 10 (52.6%) | 0.70 |
| | Largest abnormal LN (mm) | 11.8 (9.11-12) | 10.1 (8-14.8) | 0.68 |
| | LN HU max | 180 (79-1192) | 278 (156-441) | 1.00 |
| | LN max VS | 3 (2-3) | 3 (2-3) | 0.66 |
| | LN max SUV | 4.56 (2.36-6.05) | 3.37 (2.83-4.15) | 0.70 |

¶ Includes 2 clinically diagnosed and 1 Xpert-positive untreated, all during follow-up.

Values are n (%) or median (IQR). Relationship between TB outcome and baseline characteristics analysed for categorical variables by Fisher's exact test, numerical variables by Mann-Whitney t test. Bold number indicates which comparisons are significantly different ($p \leq 0.05$). BMI, body mass index; clinically diagnosed, TB symptom positive *Mtb* culture and Xpert negative; CRP, C-reactive protein; CXR, chest X-ray; ESR, erythrocyte sedimentation rate; FDG, Fluoro deoxyglucose; $HU_{max}$, maximum Hounsfield units; IFNγ, interferon-gamma; LN, lymph node; N:L, neutrophil:lymphocyte ratio in blood; N:M, neutrophil:monocyte ratio in blood; QFN+, QuantiFERON positive; $SUV_{max}$, maximum standardised uptake value; VS, visual score; WCC, whole cell count.

**Table S5. Univariate and multivariate analyses of odds of having FDG-avid lymph nodes on baseline PET/CT by main covariates. n=247 unless stated.**

| | Variable | | Crude ORs (95% CI) | Adjusted ORs (95% CI) |
|---|---|---|---|---|
| DEMOGRAPHICS | Age (years) | 18-30 | 1 | 1 |
| | | 31-65 | **3.80 (1.89-7.65)*** | **2.46 (1.16 – 5.18)*** |
| | Sex | Male | 1 | |
| | | Female | 1.26 (0.66-2.36) | |
| | Smoker ever | No | 1 | |
| | | Yes | 1.00 (0.52-1.90) | |
| | BMI | ≤25 | 1 | |
| | | >25 | 1.02 (0.54-1.90) | |
| | Previous TB | No | 1 | |
| | | Yes | 1.95 (0.87-4.33) | |
| EXPOSURE | Sleep location | Different house | 1 | |
| | | Same house | 0.94 (0.46-1.95) | |
| | | Same room | 1.59 (0.63-3.99) | |
| | Hours of contact/day | 0-6 | 1 | 1 |
| | | 7-12 | 1.86 (0.87 – 3.95) | 1.63 (0.70 – 3.78) |
| | | 13-18 | 2.78 (0.94 – 8.34) | 2.97 (0.83 – 10.68) |
| | | >18 | **4.64 (1.73 – 12.41)*** | **5.80 (1.91 – 17.79)*** |
| | Index case smear (n=190) | Neg | 1 | |
| | | 1 or 2+ | 1.33 (0.50 – 3.58) | |
| | | 3+ | 0.90 (0.37 – 2.17) | |
| INFECTION/DISEASE STATUS | QuantiFERON status (any) | NEG | 1 | 1 |
| | | POS | **4.32 (1.26 – 14.88)*** | 2.41 (0.66 – 8.76) |
| | Parenchymal abnormalities | Normal | 1 | 1 |
| | | Other lesions | 2.10 (0.88 – 4.99) | 1.99 (0.78 – 5.09) |
| | | Subclinical TB-inactive | **6.85 (2.37 – 19.72)*** | **5.78 (1.94 – 17.25)*** |
| | | Subclinical TB | **11.93 (3.82 – 35.76)*** | **9.44 (3.19 – 26.94)*** |

OR, Odds ratio; * p<0.05 ** p<0.01 ***p<0.001.
